# Novel multiplex tools in an epidemic panel improve prediction of RSV infection dynamics and disease burden – a RESPINOW analysis

**DOI:** 10.1101/2024.11.20.24317653

**Authors:** Manuela Harries, Carolina J. Klett-Tammen, Isti Rodiah, Alex Dulovic, Veronika K Jaeger, Jessica Krepel, Sebastian Contreras, Katrin Maak, Patrick Marsall, Annette Möller, Jana-Kristin Heise, Stefanie Castell, RESPINOW study group, Nicole Schneiderhan-Marra, André Karch, Berit Lange

## Abstract

Respiratory Syncytial Virus (RSV) is one of the leading causes of morbidity and mortality among infants and adult risk groups worldwide. Substantial case-underdetection and gaps in the understanding of reinfection dynamics of RSV limit reliable projection estimates.

Here, we use a novel RSV multiplex serological assay in a population-based panel to estimate season and age-specific probability of reinfection and combine it with sentinel and notification data to parameterize a mathematical model tailored to project RSV dynamics in Germany from 2020 to 2023.

Our reinfection estimates, based on a 20% post-F and a 45% N antibody increase in the assay over consecutive periods, were 5·7% (95%CI: 4·7-6·9) from 2020 to 2022 and 12·7% (95%CI: 10·5-15·2) from 2022 to 2023 in adults. In 2021, 30-39 year olds had a higher risk of reinfection, whereas in 2022, all but the 30-39 age group had an increased risk of reinfection. This suggests age-differential infection acquisition in the two seasons, e.g. due to still stronger public health measures in place in 2021 than in 2022.

Model-based projections that include the population-based reinfection estimations predicted the onset and peak for the 23/24 RSV season better than those only based on surveillance estimates.

Rapid, age-specific reinfection assessments and models incorporating this data will be critical for understanding and predicting RSV dynamics, especially with changing post-pandemic patterns and new prevention strategies e.g. monoclonal antibody.

Helmholtz Association, EU Horizon 2020 research and innovation program, Federal Ministry of Education and Research, and German Research supported this work.

## Introduction

Respiratory syncytial virus (RSV) is one of the leading causes of severe respiratory disease in infants and adults^1,2^. RSV causes only partial immunity, so that reinfections are an important part of transmission dynamics^3,4^. Typically, the primary infection in young children causes severe disease, leading to an estimated 245,244 annual hospital cases in Europe, with most of them being infants (under 1 years)^5^. Reinfections are typically not as severe. However, among the elderly (60+), they led to approximately 470,000 hospitalizations and 33,000 deaths in high-income countries in 2019^1^. Despite these high numbers, RSV has only recently been categorized as a notifiable disease in the majority of EU/EEA countries^6^, e.g. since July 2023 in Germany^7^. While this will result in an increase in publicly accessible data, without targeted efforts to correct systematic biases like health care seeking behavior, usability of this data for dynamic modelling remains limited due to age- and season differential underdetection^8^.

Changes in post-pandemic RSV patterns and the roll-out of new preventive strategies are expected to substantially affect RSV dynamics in the upcoming years.

Prior the COVID-19 pandemic, RSV infections peaked in early January in many European countries^9^. However, from 2020 to 2022, the dynamics of RSV infections and hospitalizations shifted. This change was partly influenced by the implementation of non-pharmaceutical interventions (NPIs), which impacted all respiratory pathogens^10–13^.

As novel prevention strategies two vaccines have been approved in Europe for the elderly (GSK, RSVPreF3) and pregnant women (Pfizer, PF-06928316)^14,15^; moreover, two monoclonal antibodies are licensed for (preterm) newborns and infants (under 8 months). Real-world evidence from the population-wide implementation of these prevention strategies shows high potential for reducing disease burden^16^. As a consequence, several European countries have defined implementation strategies^17–19^.

Current understanding of RSV transmission dynamics relies partly on seroprevalence studies^20^, which are often non-continuous and lack real-time and season-specific estimates. This makes them insufficient for reliable age-group projections. More importantly, transmission dynamics after early childhood and the effects of prior infections on secondary infections are mostly unknown^21^. In this study, we used data from an RSV multiplex serological assay in an infectious disease focussed population-based cohort (MuSPAD) to estimate adult age and season-specific reinfection probability. We combined this information with sentinel and RSV notification datasets to improve predictions from a mathematical model for RSV trends in Germany.

## Methods

### Study design

We re-invited MuSPAD (Multilocal and Serial Prevalence Study of Antibodies against Infectious Diseases in Germany^22^) participants from three (June-July 2022) and four (April-June 2023) regions^23^ (fig S1). Using a multiplex-based serological assay, we measured RSV IgG antibodies titres changes to RSV post-F protein from 2020 to 2023. The antigen-derived 9-plex immunoassay measures antibody titres towards post-F, Nucleoprotein (N), G proteins and subtype A or subtype B G antibodies that allowed classification as a prior infection with either subtype A or subtype B^24^. We evaluated different cut offs for reinfection definition (post-F Δ20-40%). We measured antibody titres at three intervals: baseline 2020/21, June-July 2022 and April-June 2023 representing RSV seasons 2021/2022 and 2022/2023 (figure 1).

**Figure 1:**
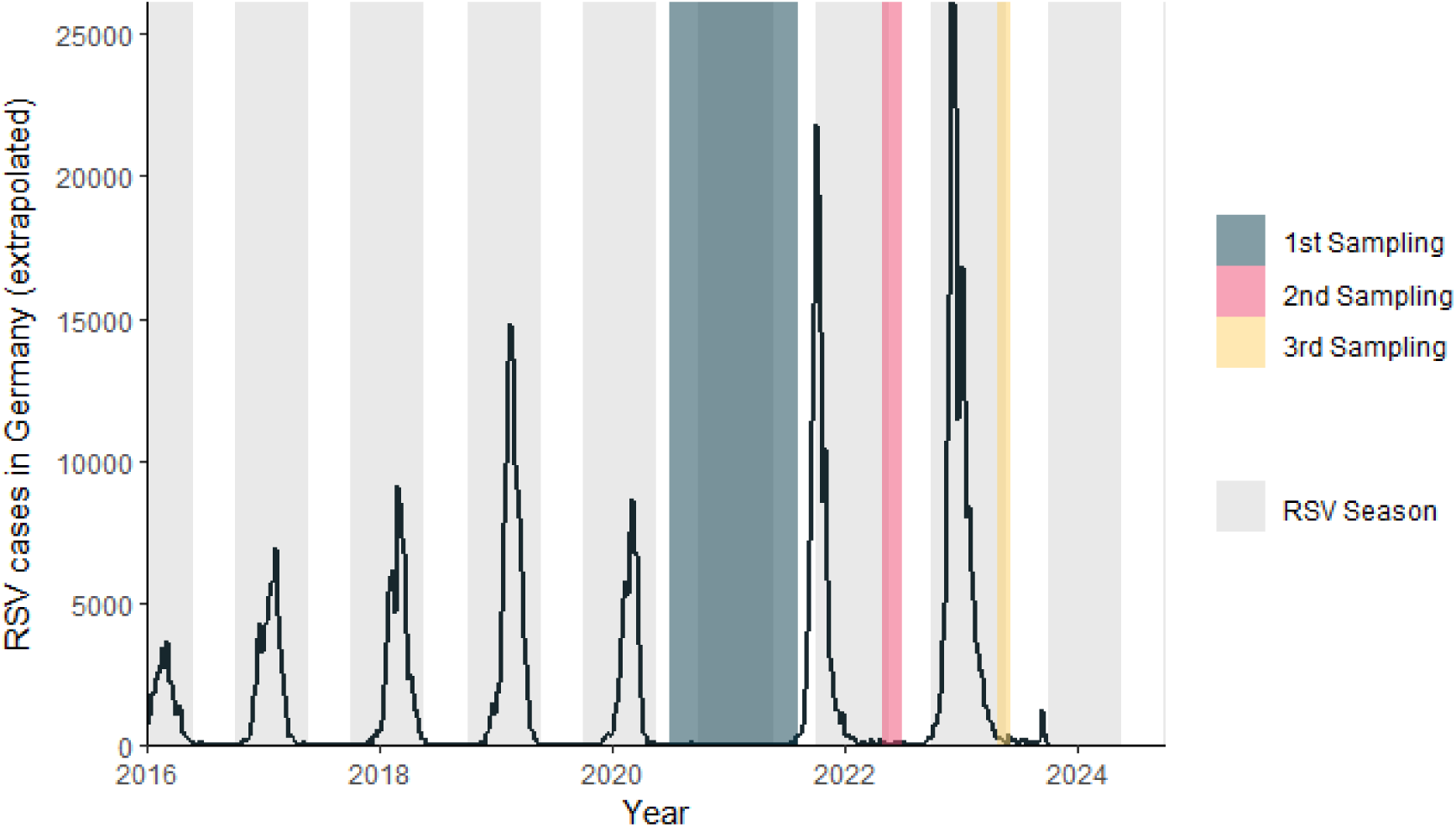
Weekly extrapolated numbers of confirmed RSV cases (notifications of RSV was limited to the state of Saxony) in Germany (light grey area RSV season CW40-20) from 2016-2023 including sampling periods of the MuSPAD participants (1^st^ sampling 2020/21 n= 32,561(dark grey shaded area), 2 sampling 2022 n=1,754 (red shaded area), 3^rd^ sampling 2023 n=2,832 (yellow shaded area). Enrolment of 836 participants with three sampling points.

We collected data on age, sex, household composition, sociodemographic, and health factors. Prior SARS-CoV-2 infection was detected by IgG antibodies against the Nucleocapsid (NC) antigen.

We provided a description of the study population and analysed Δ titre changes of two proteins (cut off value of 20% post-F and additionally 45% N-protein for quality assurance) to identify RSV reinfections in age groups (18-29, 30-39, 40-49, 50-59, 60-69, 70-79 and 80+ years). We assume Δ2020-2022 represents the 2021/22 RSV season and Δ2022-2023 the 2022/23 RSV season, since few cases were reported between 2020 and 2021.

We used logistic regression to explore how age, children under 14 in the household, and SARS-CoV-2 serology affect reinfection risk each season. We adjusted for household size, SARS-CoV-2 vaccination status, age, chronic lung disease, contacts, and children under 14 living in the household, study region, and sex/gender. Additional variables were based on peer-reviewed Directed Acyclic Graphs (Supplement DAG). Adjusted relative risks (RR) and their 95% confidence intervals (CIs) were computed from the information provided in the respective logistic model fits (Supplement).

We determined the probability of reinfection for each age group independently for 2021-2022 and 2022-2023. This probability is calculated using the two fitted logistic regression models. The statistical analyses were performed using R Version 4.0.2 and STATA/IC 14.2.

### Additional data sources

We used publicly accessible data. Details are in supplement, but in short for RSV notifications (2017-2023) we used notification data from Saxony from the public database on reportable diseases (SurvStat@RKI). We extrapolated Saxony’s age-specific data using national population statistics to estimate figures for Germany. Country-wide notification of RSV cases was only fully implemented in Germany in 2023.

We extracted weekly data on acute respiratory infections (ARI) and influenza-like illness (ILI) from 2016 to 2023 using two sources: Arbeitsgemeinschaft Influenza (AGI) and GrippeWeb. AGI is a sentinel surveillance system testing for various respiratory infections, and GrippeWeb is a citizen science platform. GrippeWeb users report regularly online ARI or ILI symptoms. We applied the positive RSV proportion from the AGI to the GrippeWeb users to estimate RSV infections for each age group, and projected this for Germany. This dataset is used in the estimation of age- and year specific underdetection factors.

For hospitalizations we used the RSV proportion of the hospital sentinel ICOSARI (ICD-10 code-based hospital surveillance of severe acute respiratory infections (SARI)) surveillance from the AGI. We extrapolated these to nationwide estimates to generate the RSV hospitalization rates for Germany.

### Age-specific SEIRS model

We specified an age-specific ODE (Ordinary Differential Equation) compartmental model to study the dynamics of RSV. The model incorporates epidemiological parameters, including age and sex/gender. We calibrated our model parameters to the measured age-specific titre changes and test different assumptions about the number of susceptible children. We considered the following compartments: immune protected (e.g. maternal or vaccinated), susceptible individuals (subdivided in classes accounting for reinfection potential), infected but not yet infectious (exposed) individuals, infectious, hospitalized, recovered, and dead. The model assumes repeated RSV reinfections increase partial immunity. In contrast to full immunity, partial immunity allows reinfection to occur at a reduced rate. We assumed that susceptible under 15 years have had up to three prior RSV infections. Those above 15 years are assumed to be susceptible individuals with four or more previous RSV infections. Table 1 outlines our assumptions of susceptibility levels to reinfection, ranging from 0% to 80% protection against reinfection.

**Table 1:** Assumption of estimated immunity level for SEIR (Susceptible-Exposed-Infected-Recovered) Model.

| Compartment | Immunity |
| --- | --- |
| $S_1$ (susceptible) | No protection |
| $S_2$ | 25% protection against reinfection in that year |
| $S_3$ | 50% protection against reinfection in that year |
| $S_4$ | 75% protection against reinfection in that year |
| $S_5$ | 80% protection against reinfection in that year but for persons older 15 years |
| P (protection, e.g. vaccinated/maternal/passive immunity) | 50% protection against reinfection in that year |

We estimated the proportion of reinfections per age group in the model using publicly available data from notifications, AGI, and SARI. We formalized the protection compartment only for those 0-1 year old due to maternal/passive immunity as immunisation strategies with vaccines and monoclonal antibodies had not yet been implemented in Germany up to 2023. Figure 2 illustrates the model structure and differential equations (table S1).

**Figure 2:**
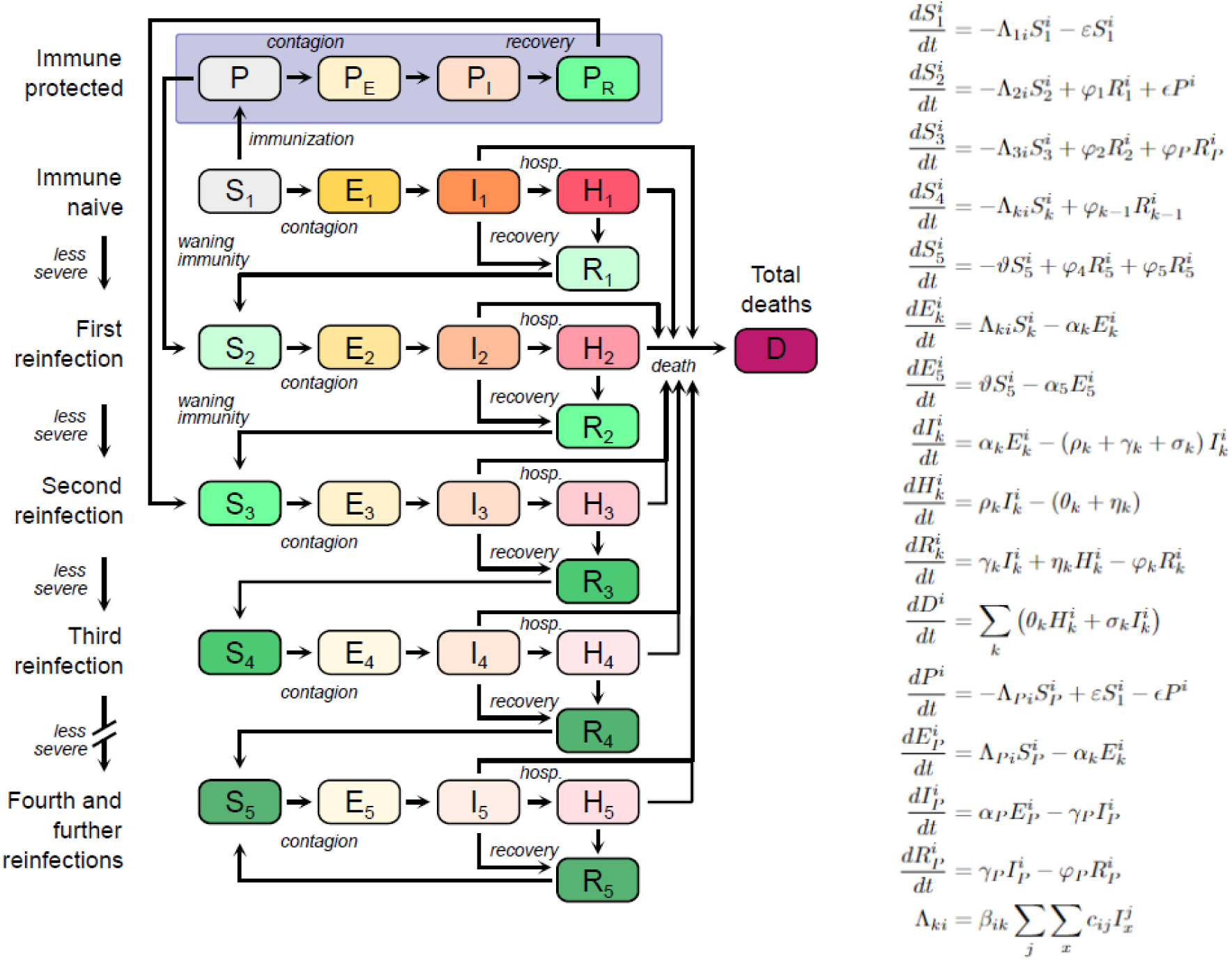
Structure and formula of the models. The compartments are classified either as susceptible (S), exposed (E), infectious (I), hospitalized (H), recovered (R), dead (D), protected (P: vaccination /passive protection), exposed after protection (P_E_), infectious after protection (P_I_), recovered after protection (P_R_). The formula includes the following components: c: Social contact rate; β: The risk of infection; α: The inverse of the incubation period (5 days); ρ: The proportion of infected individuals requiring hospitalization; γ: The inverse of the time an infected individual recovers (7 days); η: The inverse of the time a hospitalized patient recovers (4 days); ε: The proportion of susceptible individuals gaining full protection; Ill: The probability of protected individuals losing full immunity; φ: The probability of recovered individuals losing full immunity; σ: The proportion of infected individuals who will die; θ: The proportion of hospitalized patients who will die; Ill: The proportion of reinfection in adults (table S1). The detailed equations are provided in the Supplement.

We implemented this model with three RSV datasets (figure 3). For each dataset, parameter fitting using an ordinary least squares methodology was two-staged. First, the model was fitted to case data. The parameters obtained here were used as initial guess for the fit to hospitalization data.

**Figure 3:**
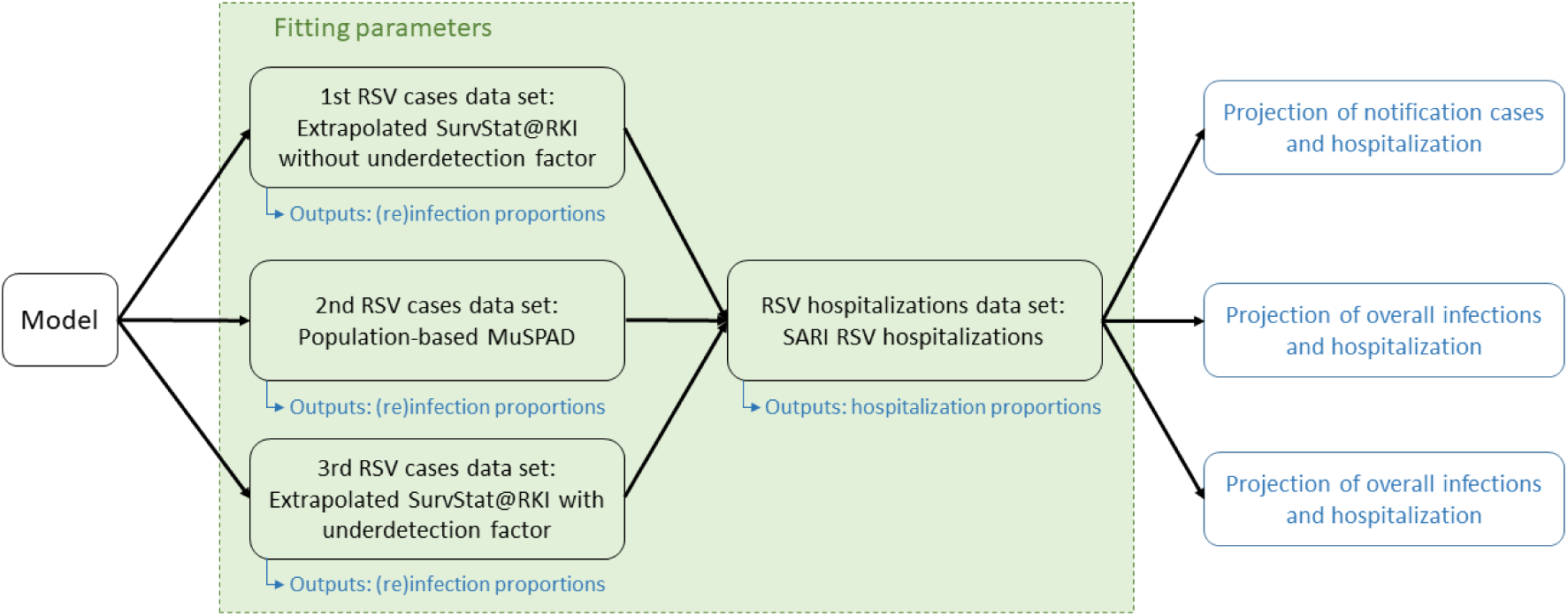
Model simulation process incorporates the integration of three RSV cases datasets and a single hospitalization dataset. The outcomes derived from the model are presented herein (blue).

#### 1. RSV notification model

The model was calibrated with extrapolated notification case data, yielding time-dependent infection parameters and reinfection proportions stratified by age groups. Subsequently, the model was fitted to extrapolated SARI hospital data of RSV specific hospitalizations to derive the corresponding hospitalization parameters from 2016 to 2022.

#### 2. RSV population-based model

The population-based model was parameterized using pre-pandemic RSV notification data, with results informing assumptions regarding the proportion of susceptible children and the incidence of reinfection in 2021 and 2022. The revised proportion of reinfection for adults was performed according to MuSPAD data for simulation parameters. We calibrated the model using the extrapolated SARI RSV hospitalization data to derive the parameters related to hospitalization.

#### 3. RSV notification model incorporating underreporting factor

For the notification model we incorporated the data with an age-specific underreporting factor. We calculated underreporting ratios by dividing the extrapolated notification numbers with our estimated case numbers for each age group (Supplement Calculation).

Using the same methodology for model fitting, we estimated model-based proportions of reinfections across age groups to Germany. We derived the hospitalization parameters by fitting the model to the SARI RSV hospitalizations.

## Results

We collected from three respectively four study cites 13,861 (Aachen, Magdeburg, Hannover and Greifswald 2021), 3,034 (Aachen, Magdeburg, Hannover 2022) and 2,832 (Aachen, Magdeburg, Hannover and Greifswald 2023) blood samples. Among these, there was an overlap of 1,754 individuals with samples from both 2020/21 and 2022, and 836 individuals with samples from 2020/21, 2022, and 2023. The median age was 55 years (IQR 40-67) in 2022 (2023: 56 year IQR 41-68) and 60% (2023: 61%) were female (table 2). Among the participants who experienced reinfection in 2021/22, 40% (40/100) have had a previous COVID-19. Subtype analysis using multiplex serology showed that among the 106 participants reinfected in 2023, of whom 11 (10·4%) were subtype A, 55 (51·9%) subtype B, and 40 (37·7%) were unclear.

**Table 2:**
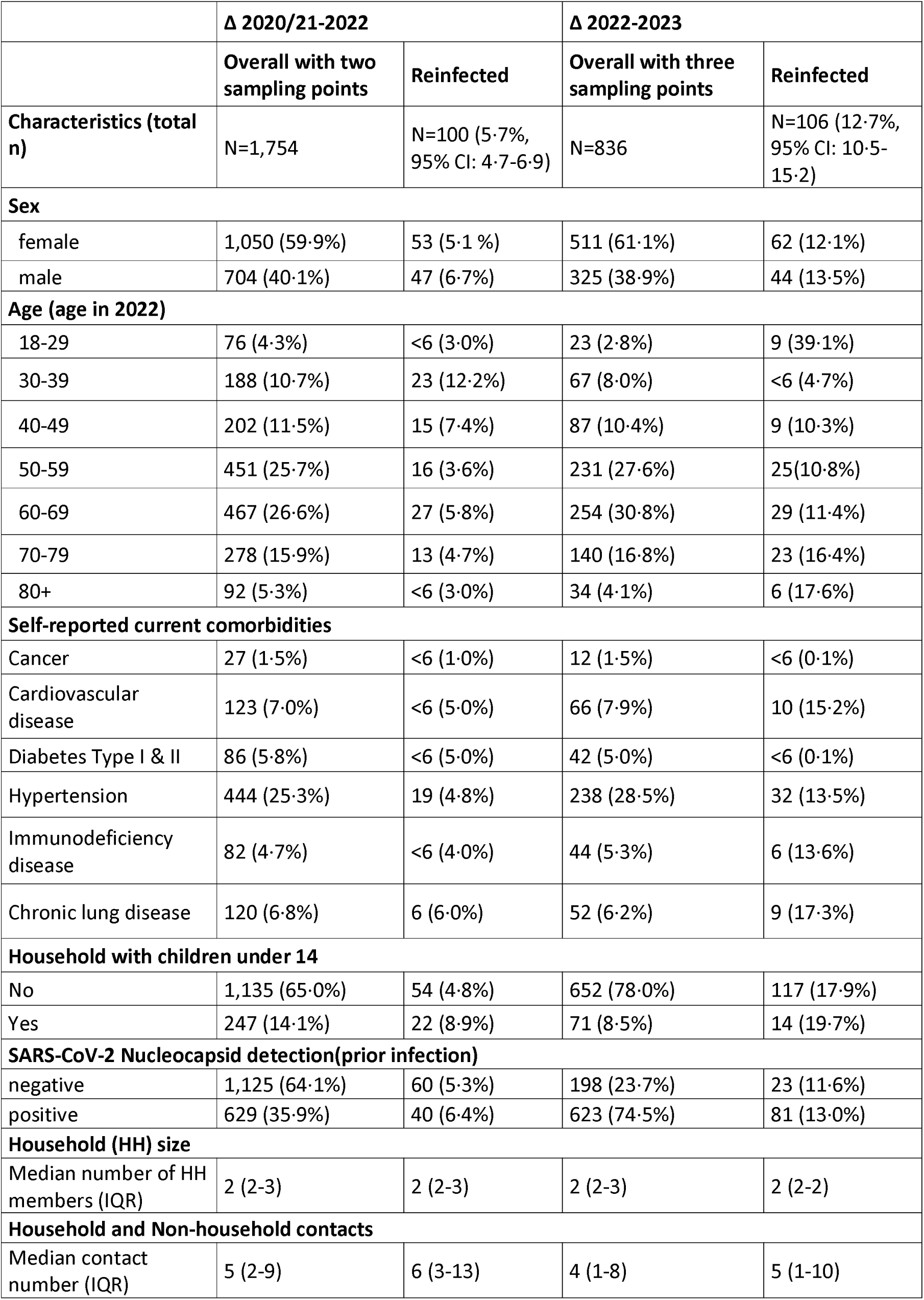
MuSPAD participant characteristics stratified by RSV reinfection status; sampling in season Δ 2021/2022 and Δ 2022/2023; Reinfection is defined as a titre increase of 20% in post-F-protein and 45% in N-protein between two sampling points (full overview table S1)

### RSV Reinfection by age groups

We use three adjustment sets accounting for prior SARS-CoV-2 infection (NC), children <14 years in household and age. The probability of reinfection is higher for all age groups in season 2022-23 compared to 2021-22 (figure 4). Within season 2022-23, the youngest age group shows the highest probability, followed by the oldest age group (figure 4a). Predicted probabilities for positive and negative SARS-CoV-2 NC antibody responses remain unchanged within the age groups (figure 3b). Having children under 14 in the household increases the predicted probability of reinfection for the 2022/23 season (figure 4c). The age group 30-39 years exhibited a higher adjusted risk ratio (aRR; Supplement DAG) during 2021-2022 season (aRR: 3·11 95%CI 0·96-10·07) compared to 2022-2023 season (aRR: 0·21 95%CI 0·08-0·56) (figure 4d, table S2).

**Figure 4:**
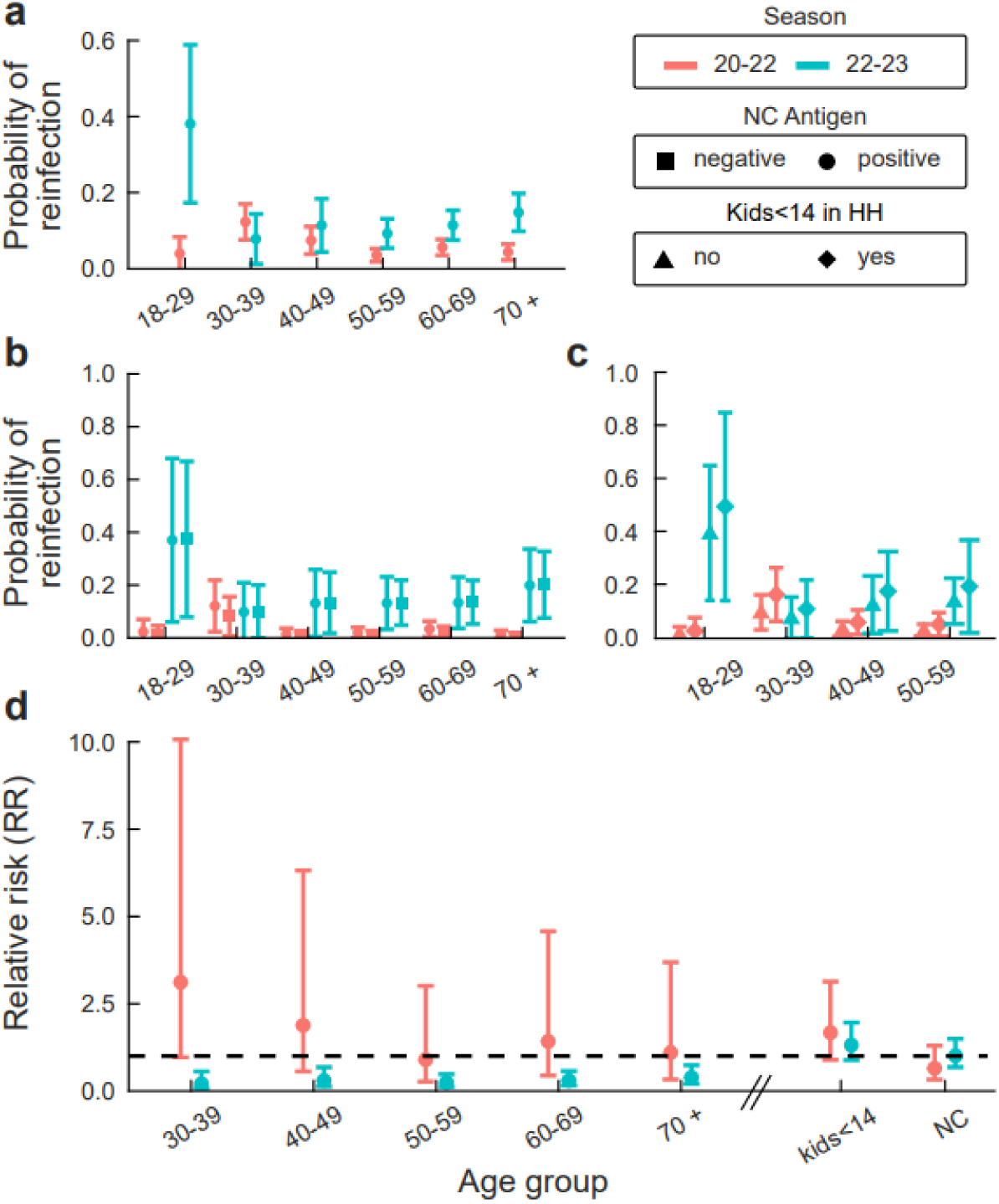
a) Predicted probability of reinfection over age groups regarding RSV season 2020/21-2022 (red dots) and 2022-2023 (blue dots; upper panel); as b) exposure SARS-CoV-2 prior infection (Nucleocapsid (NC); positive NC=circle and negative NC=square; middle left panel) and c) exposure children under 14 years living in the household (HH) no=triangle and yes=diamante; middle right panel) based on three different models and their adjustment sets. d) Adjusted relative risk (RR=1 (black dashed line; bottom panel) for adjustment set see Supplement DAG)

The number of household contacts (figure S2) increased the likelihood of having a reinfection in 2020/21-2022 (OR 4·22; 95%CI 0·61-22·20), but not in the 2022/2023 season. The number of non-household contacts did not have an effect on the odds ratios (OR 0·98; 95%CI 0·89-1·03).

### Model-based RSV reinfection proportions

We fitted the parameters of our model for RSV using extrapolated data for weekly reports in Germany. With this we estimated the weekly RSV reinfection proportions for pre- and post-pandemic seasons by dividing the total infections predicted by our model by the overall population for each season. Model-based reinfection estimates were consistently lower than population-study based reinfection estimates. In pre-pandemic seasons, the pattern and proportion of RSV reinfections in each age group were similar, ranging from 0% to 1·2% (figure 5a). In the 2021/22 season, there was a drop in reinfections, while 2022/23 we saw a sharp increase to 10% in the oldest age group (figure 5b). The mean trends on RSV reinfection consistently mark the importance of the 15-34 age group, leading the dynamics (figure 5c).

**Figure 5:**
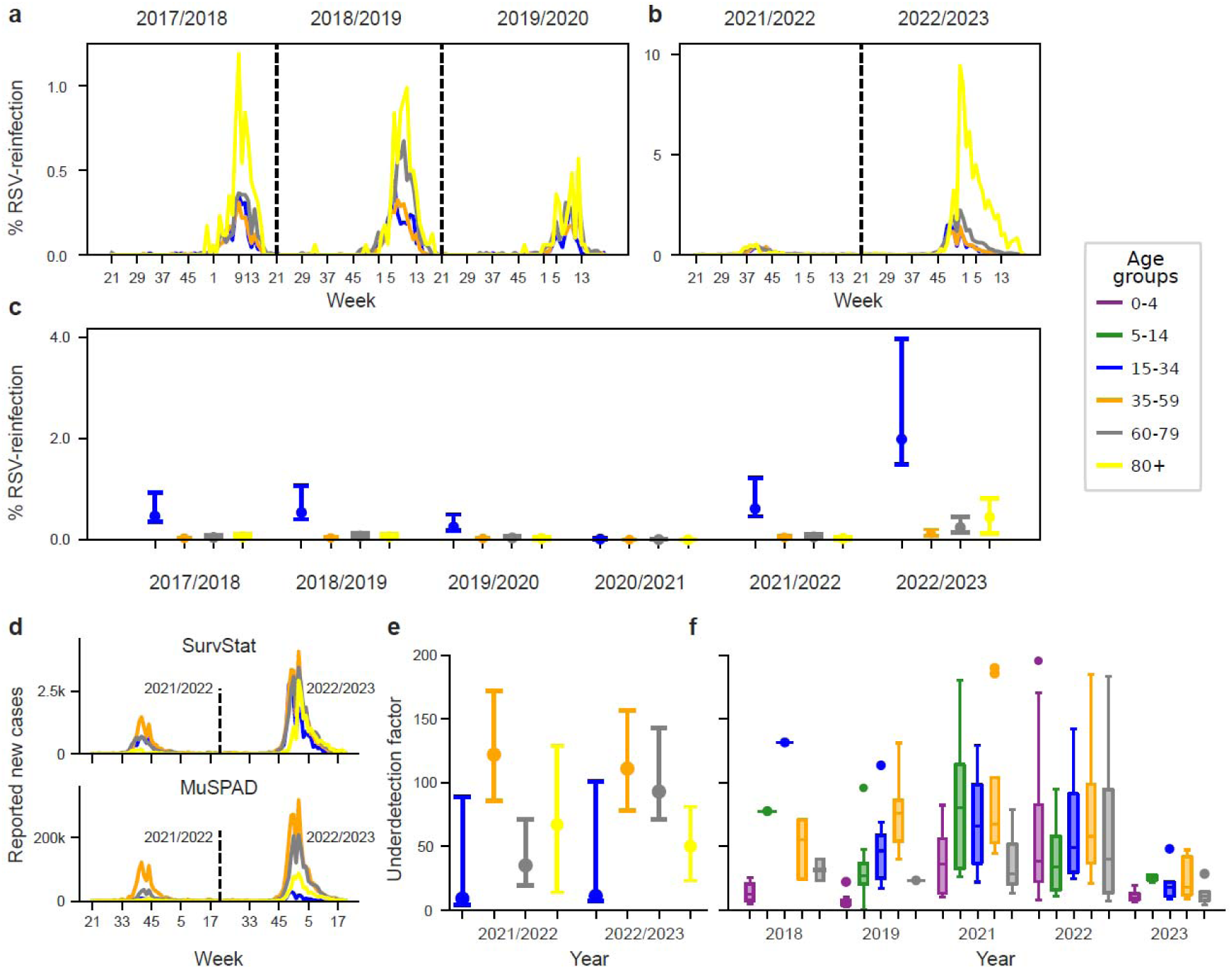
Estimated and extrapolated weekly proportion of RSV reinfection by age group based on SurvStat@RKI data over calendar weeks of (a) 2017-2020 and (b) 2021-2023. (c) Estimated proportion of RSV reinfection extrapolated by age group based on SurvStat@RKI data from 2017-2023. (d) Estimated RSV infections by age group based on SurvStat@RKI data and population-based MuSPAD data for the seasons 2021/2022 and 2022/2023 by age group and calendar week. Please be aware of the different scales of the y-axis. (e) Underdetection factor for RSV reinfection by age group, estimated by comparing the RSV reinfections in MuSPAD with SurvStat@RKI. (f) Underdetection factor of extrapolated reported cases, defined by calculated cases using GrippeWeb and ARI surveillance stratified by age group over the 2018-2023. Unexplained outliers were deleted to ensure validity.

By comparing the proportion of RSV reinfection from the MuSPAD data and the estimated new notification cases based on SurvStat@RKI, we calculated an underdetection factor for adult RSV reinfection by adult age group for seasons 2021/22 and 2022/23 (figure 5e), with high underdetection seen in the 15-34 age group. T (figure 5f).

### Projection of RSV cases and hospitalizations

Using SurvStat@RKI data from only Saxony without an underdetection factor, we predicted extrapolated RSV cases for 2023/2024 (figure 6a). We also used population-based data (figure 6b) and SurvStat@RKI data with an estimated underdetection factor (figure 6c) for overall predictions. Although the onset matches previous predictions, predicted cases, except for those 80+, are higher due to the underdetection factor in population-based data.

**Figure 6:**
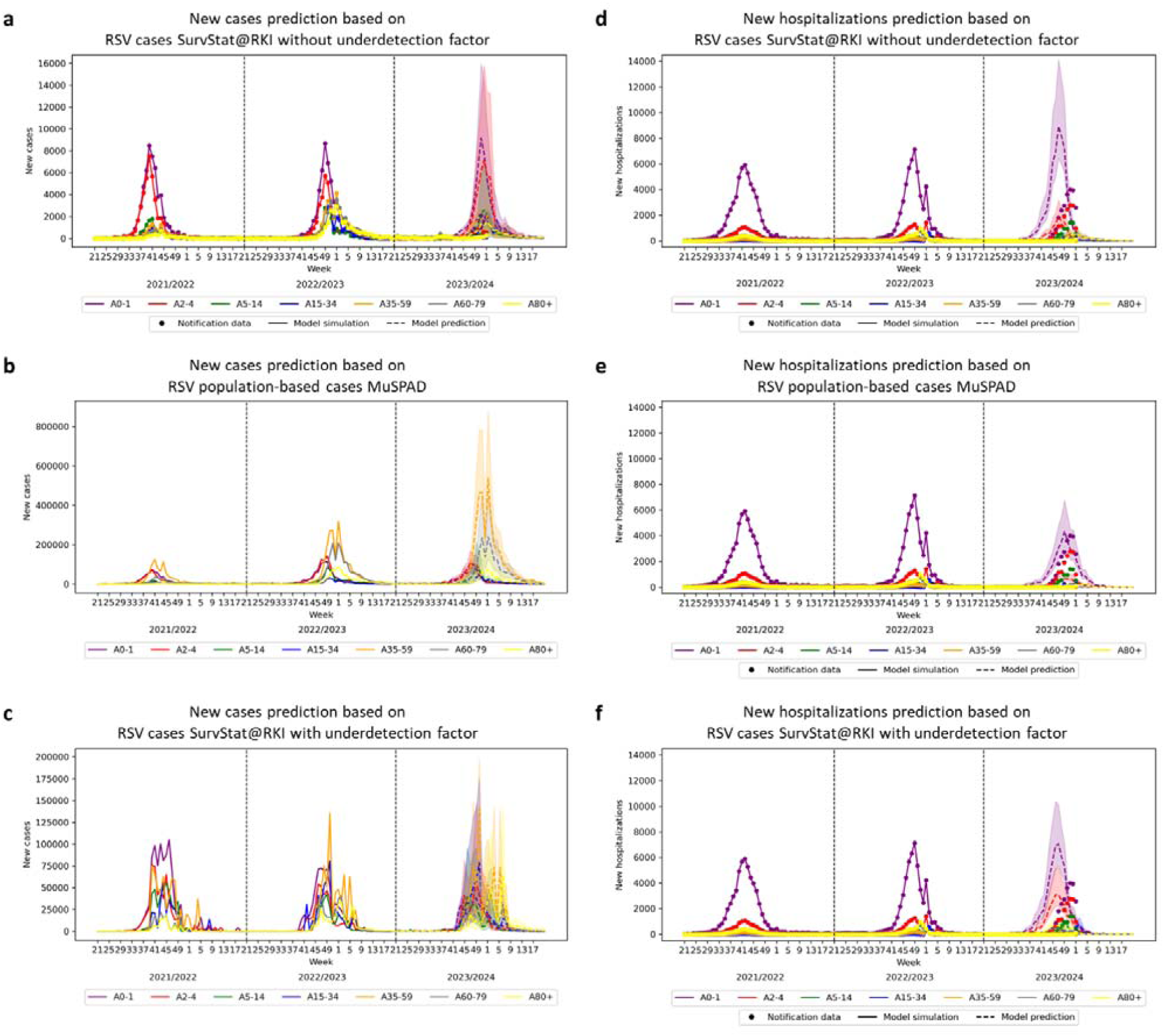
(a) RSV reported (points), model simulated (solid line) and predicted (dashed line) new cases by age group based on SurvStat@RKI data without underdetection factor and 95%CI (shaded area). (b) RSV model simulated (solid line) and predicted (dashed line) new cases by age group based on MuSPAD data with 95%CI (shaded area). (c) RSV model simulated (solid line) and predicted (dashed line) new cases for 2023/2024 by age group based on SurvStat@RKI with underdetection factor from sentinel data with 95%CI (shaded area). (d) Prediction (points) of new RSV hospitalizations for 2023/2024 by age group based on infected cases SurvStat@RKI data without underdetection. (e) Prediction of new RSV hospitalizations for 2023/2024 by age group based on MuSPAD data. (f) Prediction of new RSV hospitalizations for 2023/2024 by age group based on from SurvStat@RKI data with underdetection factor from sentinel (AGI and GrippeWeb) data.

We predicted new RSV hospitalizations using SARI hospitalization data and three sources: SurvStat@RKI without underdetection factor (figure 6d), population-based (MuSPAD (figure 6e), and SurvStat@RKI with underdetection factor (figure 6f). The peaks in RSV hospitalizations are predicted from week 45 to 49 for season 2022/23 and 2023/24. The model with population-based data predicts fewer new RSV hospitalizations than SurvStat@RKI data, regardless of the underdetection factor. The inclusion of the underdetection factor in SurvStat@RKI data shows higher hospitalization numbers, especially in the 2-4 years age group, compared to data without it (figure 6d; figure 6f). However, for the 0-1 year age group, the peak in new RSV hospitalizations is slightly lower with underdetection than without it.

Regarding the prediction of new hospitalizations for the 2023/2024 season, there is a peak delay of about four weeks. Overall the prediction using population-based reinfection estimates is closer in all age-groups to the actual trajectory of cases than the surveillance based predictions. In the 0-1 year age group, the peak predictions in the MuSPAD data (n=4,326) is almost as close as the absolute numbers seen in the SARI RSV hospitalization data (n=3,994). In addition, when compared to the previous two seasons, there was a notable increase in the number of RSV hospitalizations for children aged 2-4 years. The peak predictions in the SurvStat@RKI data with an underdetection factor of 2-4 years of age is nearly the same as the absolute numbers seen in the SARI RSV hospitalization data.

## Discussion

In this work, we used longitudinal serological population-based MuSPAD data and publicly available sentinel and notification data to parametrize an RSV ODE model to project the RSV dynamics in Germany for the 2023/24 season. We published these projections in a common statement by the modelling network for severe infectious diseases MONID in 2023^25^, and evaluated there accuracy here, retrospectively. We show a qualitatively adequate forecast with minor deviations from the actual season, in particular using the reinfection estimators provided by serological indicators from the population-based cohort. This lays a necessary foundation for future efforts to model RSV, including with incoming prevention strategies and underscores the necessity of using estimates from population cohorts combined with population-scalable multiplex serological assays.

Our results indicate that the dynamics of RSV reinfections in the adult population are season specific. While in the 2021/22 season reinfections estimates were lower in most age groups apart from the 30-39 year olds than in pre-pandemic periods they were higher in the 2022/23 season^26^. Young adults (30-39 years) were more likely to be reinfected in 2021/22, and there was a tendency for an association of reinfection risk with having children (<14y) in the household were evident from 2020-2022, but not during 2022-2023.

Population-based reinfection estimates for 2021/2022 and 2022/2023 respiratory season help investigate post-pandemic trends, which are not obvious from sentinel and notification data in Germany. Cai et al.^27^ have described the characteristics of these seasons for children, highlighting how they differ from average pre-pandemic seasons. They show that in both seasons hospitalizations for children over two were 3-4 times higher than pre-pandemic levels (1·5-2 times in those <2 years). Our cohort does not include children. However, the higher number of affected older children align with our finding of a higher-than-usual proportion of reinfected adults aged of 30-39 who are likely the parents of these children. This reflects the dynamic effects of an increased infection frequency in the population. In addition to different RSV subtypes being responsible for the seasons (A in 21/22 and B in 22/23), this suggests differential infection acquisition in the two seasons. In the 21/22 season infections were more likely acquired via children in the household whereas in the 22/23 season this was more likely via non-household contacts or adult family members. This agrees well with our findings that in the 21/22 season the number of household contacts was associated with reinfection risk.

Comparing reinfection estimates from population-based data with surveillance data we found a mean underdetection factor of 33·8 for RSV infection in the notification data in Germany from 2018 and 2023, although we used a conservative approach to minimise false positive classification, meaning that the reinfection might be even higher. Expectedly, in line with disease severity risk, we found underdetection to be especially high in the 35-59 and 15-34 age groups.

Our ODE model effectively captured dynamic patterns in previous seasons and, when combined with population-based surveillance data, predicted for 2023/2024 a similar pattern as in 2022. Interestingly, the simulation taking into account population-based estimates also yielded results closer to actual dynamics in those age-groups (the children) where no population-based estimates were available. While estimates for this age group would enhance accuracy, this emphasizes the need for estimates of age groups not typically at risk for severe disease^28^. We integrated protection compartments to our model able to mirror the prevention strategies currently being implemented. To assess these a thorough comprehension of the current RSV dynamics at the population level is necessary.

Our study has limitations. First, although we demonstrated the robustness of our results with regard to the thresholds in post-F and N-protein antibodies there is no gold-standard available for this yet. Although a less stringent cut-off has been used in previous studies in specific sub-groups^29^, our sensitivity analyses showed more reliable results with this more conservative approach. The duration between infection and the evaluation of the post-F-protein varied among the participants. Here, due to a lack of previous evidence we did not include assumptions for titre waning in our calculations, although this data was included as part of the RSV immunoassay validation^24^. We also did not include assumptions for the proportion of same-season reinfections and did not investigate those persons (4) with serological indication of reinfection in both season further. And while we found no relation between previous SARS-CoV-2 infection and RSV reinfection, we did not assess disease severity, so our findings do not contradict findings of other groups showing a higher risk of notified or clinically apparent RSV infections in those with (recent) SARS-CoV-2 infection.

The nationwide mandatory RSV reporting system in Germany from 2023 onwards will have similar detection biases as the previous system in Saxony. Physicians due to the lack of a therapeutic consequence do not normally test patients with mild to moderate ARI or ILI symptoms. Also, not everyone with such symptoms visits a physician, and not all RSV cases show symptoms. Therefore, our approach combining the surveillance system with population-based and additional data sources aids in monitoring the effectiveness of prevention strategies. This would be in particular beneficial in combination with in-depth ARI sentinel systems from day-cares and paediatricians of single federal states, as already in place in some European regions^30^. With post-pandemic dynamic changes and incoming prevention strategies, appropriate capacities must now be established to provide model-usable data for accurate projections, build up models able to integrate these data and with that evaluate continuously the effectiveness of prevention strategies so that these may be adapted. We plan to integrate our 2023/24 data projections into MONID and publish them on www.respinow.de as soon as possible.

## Data Availability

All data produced in the present study are available upon reasonable request to the authors

https://serohub.net/

## Acknowledgements

We sincerely thank all the participants from Aachen, Hannover, Magdeburg and Greifswald for endorsing our study. We are grateful to Kim Busch and Sabine Pape for answering the participant’s questions and concerns. We thank our colleagues in Osnabrück laboratories for the laboratory work. We thank Daniela Gornyk and Pilar Hernandez to have built the initial study population (MuSPAD), and Julia Ortmann and Max Hassenstein for the initial data cleaning, and Daniel Alexander Schulze for study assistance and the whole MuSPAD study group (Stefanie Castell, Pilar Hernandez, Monike Schlüter, Gerhard Bojara, Kristin Frank, Knut Grubbe, Torsten Tronn, Oliver Kappert, Winfried Kern, Thomas Illig, Norman Klopp, Gottfried Roller, Michael Ziemons, Daniela Gornyk, Manuela Harries, Stephan Glöckner, Jana-Kristin Heise, Yvonne Kemmling, Barbora Kessel, Gérard Krause, Berit Lange, Henrike Maaß, Julia Ortmann, Monika Strengert, Tobias Kerrines).

## Data sharing statement

We will share anonymized data utilized in this study with other academic researchers upon request. The variables can include study site information, assay information, bio sample type, demographic information, and test results. For further information, please contact. Additionally, all links to publicly available data sources are in the supplement.

### Authors’ contributions

Conceptualization: B.L.

Data quality: A.M., J.K., M.H., CJ.KT., MA.K., V.J.;

Formal analyses: A.M., CJ.KT., M.H., I.R., J.K.;

Visualization: A.M., I.R., CJ.KT., M.H., J.K. S.C;

Project administration: M.H., B.L., CJ.KT;

Laboratory analysis: A.D., P.M, N.S.;

Writing-original draft: M.H., CJ.KT., B.L., I.R. J.K. A.M. S.C.;

Funding acquisition: B.L., A.K., N.S, St Ca;

Writing-review & editing: CJ.KT., M.H., I.R., J.K., A.M.,V.J., MA.K, B.L., A.K., P.M., A.D., N.S., S.C. St Ca.; JK.H. RESPINOW study group;

All authors have read and agreed to the published version of the manuscript.

## References

1. Savic M, Penders Y, Shi T, Branche A, Pircon JY. Respiratory syncytial virus disease burden in adults aged 60 years and older in high-income countries: A systematic literature review and meta-analysis. Influenza Other Respir Viruses 2023; 17(1): e13031.

2. Ambrosch A, Luber D, Klawonn F, Kabesch M. Focusing on severe infections with the respiratory syncytial virus (RSV) in adults: Risk factors, symptomatology and clinical course compared to influenza A / B and the original SARS-CoV-2 strain. J Clin Virol 2023; 161: 105399.

3. Bont L, Versteegh J, Swelsen WT, et al. Natural reinfection with respiratory syncytial virus does not boost virus-specific T-cell immunity. Pediatric research 2002; 52(3): 363–7.

4. Hall CB, Walsh EE, Long CE, Schnabel KC. Immunity to and frequency of reinfection with respiratory syncytial virus. Journal of Infectious Diseases 1991; 163(4): 693–8.

5. Del Riccio M, Spreeuwenberg P, Osei-Yeboah R, et al. Defining the Burden of Disease of RSV in the European Union: estimates of RSV-associated hospitalisations in children under 5 years of age. A systematic review and modelling study. J Infect Dis 2023.

6. Control ECfDPa. Intensified circulation of respiratory syncytial virus (RSV) and associated hospital burden in the EU/EEA. 2023.

7. Justiz) BBd. Infektionsschutzgesetz - IfSG §7. 2023.

8. Teirlinck AC, Johannesen CK, Broberg EK, et al. New perspectives on respiratory syncytial virus surveillance at the national level: lessons from the COVID-19 pandemic. Eur Respir J 2023; 61(4).

9. Robert-Koch-Institut. RSV notifications for Saxony. 2021 (accessed accessed 09/2021.

10. Koltai M, Krauer F, Hodgson D, et al. Determinants of RSV epidemiology following suppression through pandemic contact restrictions. Epidemics 2022; 40: 100614.

11. Juhn YJ, Wi CI, Takahashi PY, et al. Incidence of Respiratory Syncytial Virus Infection in Older Adults Before and During the COVID-19 Pandemic. JAMA Netw Open 2023; 6(1): e2250634.

12. Eden JS, Sikazwe C, Xie R, et al. Off-season RSV epidemics in Australia after easing of COVID-19 restrictions. Nat Commun 2022; 13(1): 2884.

13. Fricke LM, Glöckner S, Dreier M, Lange B. Impact of non-pharmaceutical interventions targeted at COVID-19 pandemic on influenza burden - a systematic review. J Infect 2021; 82(1): 1–35.

14. EMA. Arexvy. Recombinant respiratory syncytial virus pre-fusion F protein, adjuvanted with AS01E. 2023. https://www.ema.europa.eu/en/medicines/human/EPAR/arexvy (accessed 05.09.2023.

15. Kampmann B, Madhi SA, Munjal I, et al. Bivalent Prefusion F Vaccine in Pregnancy to Prevent RSV Illness in Infants. N Engl J Med 2023; 388(16): 1451–64.

16. Martinón-Torres F, Mirás-Carballal S, Durán-Parrondo C. Early lessons from the implementation of universal respiratory syncytial virus prophylaxis in infants with long-acting monoclonal antibodies, Galicia, Spain, September and October 2023. Eurosurveillance 2023; 28(49): 2300606.

17. ECDC. Vaccine Scheduler: RSV: Recommended vaccinations. 2023. https://vaccine-schedule.ecdc.europa.eu/Scheduler/ByDisease?SelectedDiseaseId=53&SelectedCountryIdByDisease=-1 (accessed 27.05 2024).

18. Bundesminsterium Soziales G, Plege und Konsumentenschutz. Impfplan Österreich 2023/2024. 2024. https://www.sozialministerium.at/Themen/Gesundheit/Impfen/Impfplan-%C3%96sterreich.html (accessed 27.05 2024).

19. Council SH. Vaccination against RSV (adults). Brussels, 2023.

20. Nakajo K, Nishiura H. Age-dependent risk of respiratory syncytial virus infection: A systematic review and hazard modeling from serological data. J Infect Dis 2023.

21. Kaler J, Hussain A, Patel K, Hernandez T, Ray S. Respiratory Syncytial Virus: A Comprehensive Review of Transmission, Pathophysiology, and Manifestation. Cureus 2023; 15(3): e36342.

22. Gornyk D, Harries M, Glockner S, et al. SARS-CoV-2 Seroprevalence in Germany. Dtsch Arztebl Int 2021; 118(48): 824–31.

23. Harries M, Jaeger VK, Rodiah I, et al. Bridging the gap-estimation of 2022/2023 SARS-CoV-2 healthcare burden in Germany based on multidimensional data from a rapid epidemic panel. International Journal of Infectious Diseases 2024; 139: 50–8.

24. Marsall P, Fandrich M, Griesbaum J, et al. Development and validation of a respiratory syncytial virus multiplex immunoassay. Infection 2024; 52(2): 597–609.

25. Calero Valdez AC, Tim; Greiner, Wolfgang; Karch, André; Kheifetz, Yuri; Kirsten, Holger; Kretzschmar, Mirjam; Kühn, Martin; Kuhlmann, Alexander; Lange, Berit; Leithäuser, Neele; Mikolajczyk, Rafael; Mohring, Jan; Nagel, Kai; Priesemann, Viola; Rehmann, Jakob; Rodiah, Isti; Scholz, Markus; Schütte, Christof; Steinmann, Maren. Szenarien zur Belastung der Bevölkerung und des Gesundheitswesens durch SARS-CoV-2-Infektionen im Winter 2023/24 und Einschätzungen zur Belastung durch andere Atemwegsinfektionen. Zenodo 2023.

26. van Summeren J, Meijer A, Aspelund G, et al. Low levels of respiratory syncytial virus activity in Europe during the 2020/21 season: what can we expect in the coming summer and autumn/winter? Eurosurveillance 2021; 26(29): 2100639.

27. Cai W, Köndgen S, Tolksdorf K, et al. Atypical age distribution and high disease severity in children with RSV infections during two irregular epidemic seasons throughout the COVID-19 pandemic, Germany, 2021 to 2023. Eurosurveillance 2024; 29(13): 2300465.

28. Rodiah I, Vanella P, Kuhlmann A, et al. Age-specific contribution of contacts to transmission of SARS-CoV-2 in Germany. European Journal of Epidemiology 2023; 38(1): 39–58.

29. Capella C, Chaiwatpongsakorn S, Gorrell E, et al. Prefusion F, postfusion F, G antibodies, and disease severity in infants and young children with acute respiratory syncytial virus infection. The Journal of infectious diseases 2017; 216(11): 1398–406.

30. NLGA. [Respiratory diseases /Influenza] Atemwegserkrankungen / Influenza. 2023. https://www.nlga.niedersachsen.de/are/uebersicht-205132.html.

