## supplement for "Novel multiplex tools in an epidemic panel improve prediction of RSV infection dynamics and disease burden – a RESPINOW analysis"

Supporting Information

Content

[Figure S3: DAG for kids in HH (<14y) and RSV reinfection risk	8](#_Toc182292028)

List of table

### 1. MuSPAD sampling/survey period for RESPINOW

The blood-sampling period took place from June 6th, 2022 to July 3rd, 2022. We chose three existing MuSPAD study sites: City Region Aachen (around 560,000 inhabitants), Magdeburg (around 236,000 inhabitants) and Hannover (around 536,000 inhabitants). From June 2022 on, we send out our online and paper-based survey to all established MuSPAD participants (Freiburg, Reutlingen, Chemnitz, Greifswald, Magdeburg, Hannover, Aachen, and Osnabrück). The blood-sampling period in 2023 took place from April to June. We added one study city (County Vorpommern-Greifswald about 237,355 inhabitants).

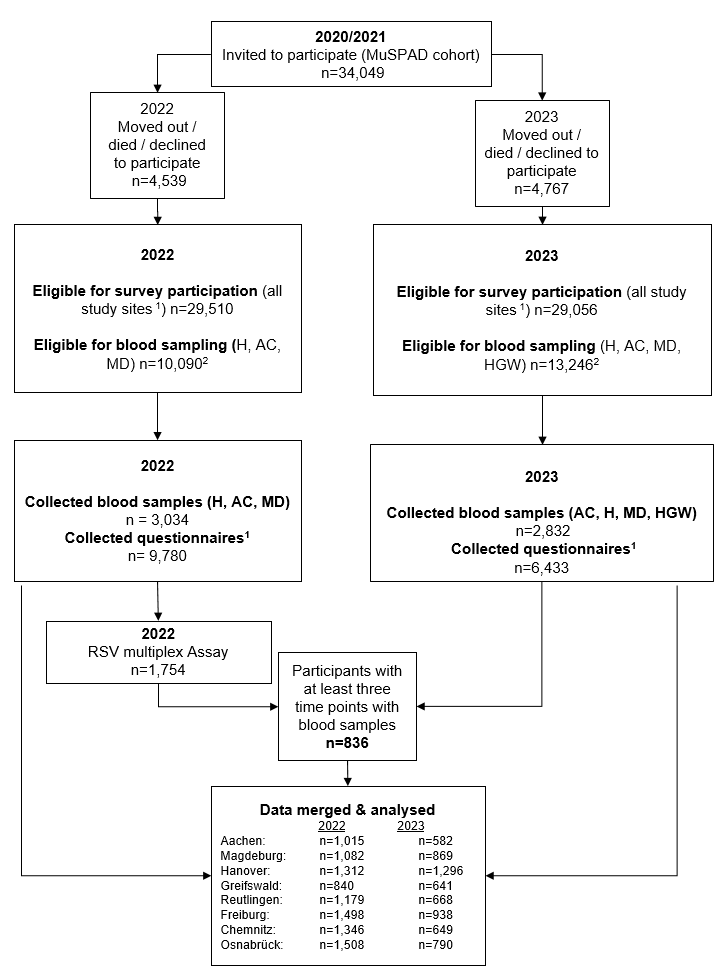

Figure 1: Flow chart of MuSPAD population and samples analysed in the seroprevalence study. Samples were analysed for RSV antibody titres towards the post-F, Nucleoprotein and a diverse mix of G proteins

^1^ Invited study locations for answering the questionnaire were Reutlingen (RT), Freiburg (FR), Osnabrueck (OS), Magdeburg (MD), Chemnitz (C)), Greifswald (HGW); Hannover( H), Aachen (AC)) ^2^ Selected study sites were invited to donate blood samples: Magdeburg (MD), Hannover( H), Aachen (AC); Greifswald (HGW) only in 2023.

Table 1: Full overview MuSPAD participant characteristics stratified by RSV reinfection status

|  | **∆ 2020/21-2022** | | **∆ 2022-2023** | |
| --- | --- | --- | --- | --- |
|  | **Overall with two sampling points** | **Reinfected** | **Overall with three sampling points** | **Reinfected** |
| **Characteristics (total n)** | N=1754 | N=100 (5·7%, 95% CI: 4·7-6·9) | N=836 | N=106 (12·7%, 95% CI: 10·5-15·2) |
| **Sex** | | | | |
| female | 1050 (59·9%) | 53 (5·1 %) | 511 (61·1%) | 62 (12·1%) |
| male | 704 (40·1%) | 47 (6·7%) | 325 (38·9%) | 44 (13·5%) |
| **Age (age in 2022)** | | | | |
| 18-29 | 76 (4·3%) | <6 (3·0%) | 23 (2·8%) | 9 (39·1%) |
| 30-39 | 188 (10·7%) | 23 (12·2%) | 67 (8·0%) | <6 (4·7%) |
| 40-49 | 202 (11·5%) | 15 (7·4%) | 87 (10·4%) | 9 (10·3%) |
| 50-59 | 451 (25·7%) | 16 (3·6%) | 231 (27·6%) | 25(10·8%) |
| 60-69 | 467 (26·6%) | 27 (5·8%) | 254 (30·8%) | 29 (11·4%) |
| 70-79 | 278 (15·9%) | 13 (4·7%) | 140 (16·8%) | 23 (16·4%) |
| 80+ | 92 (5·3%) | <6 (3·0%) | 34 (4·1%) | 6 (17·6%) |
| **Study location** | | | | |
| Aachen | 558 (31·8%) | 26 (4·7%) | 203 (24·3%) | 27 (15·3%) |
| Hanover | 575 (32·8%) | 30 (5·2%) | 335 (40·1%) | 38 (12·8%) |
| Magdeburg | 621 (35·4%) | 44 (7·1%) | 298 (35·7%) | 41 (16·0%) |
| **Comorbidities present** | | | | |
| Cancer | 27 (1·5%) | <6 (1·0%) | 12 (1·5%) | <6 (0·1%) |
| Cardiovascular | 123 (7·0%) | <6 (5·0%) | 66 (7·9%) | 10 (15·2%) |
| Diabetes | 86 (5·8%) | <6 (5·0%) | 42 (5·0%) | <6 (0·1%) |
| Hypertension | 444 (25·3%) | 19 (4·8%) | 238 (28·5%) | 32 (13·5%) |
| Immunosuppression disease | 82 (4·7%) | <6 (4·0%) | 44 (5·3%) | 6 (13·6%) |
| Chronic lung disease | 120 (6·8%) | 6 (6·0%) | 52 (6·2%) | 9 (17·3%) |
| **Smoking status** | | | | |
| Daily smoker | 154 (8·8%) | 6 (6·0%) | 60 (7·2%) | 7 (11·7%) |
| Occasional smoker | 65 (3·7%) | <6 (3·0%) | 18 (2·2%) | <6 (2·8%) |
| Previous smoker | 531 (30·3%) | 28 (5·3%) | 255 (30·0%) | 38 (14·9%) |
| Never smoker | 1000 (57·0%) | 62 (6·2%) | 502 (60·1%) | 58 (11·6%) |
| **Education** | | | | |
| Higher education | 1131 (64·5%) | 66 (5·8%) | 522 (62·4%) | 60 (11·5%) |
| Certification after 10 years | 483 (28·0%) | 30 (6·2%) | 252 (30·4%) | 40 (15·9%) |
| Certification after 9 years | 125 (7·1%) | <6 (3·0) | 58 (6·9%) | 6 (10·3%) |
| No certification | <6 (0·2%) | 0 (0·0%) | <6 (0·1%) | <6 (0·0%) |
| I don´t want to answer | <6 (0·3%) | <6 (0·0%) | <6 (0·2%) | <6 0·0%) |
| Missing | 7 (0·5%) | <6 (0·1%) | <6 (0·1%) | <6 (0·0%) |
| **Employment status** | | | | |
| Unemployed | 25 (1·4%) | <6 (0·0%) | 13 (1·6%) | <6 (0·9%) |
| Student | 48 (2·7%) | <6 (3·0%) | 14 (1·7%) | 6 (42·9%) |
| Teaching sector | 81 (4·6%) | <6 (5·0%) | 34 (4·1%) | 5 (14·7%) |
| Medical sector | 100 (5·7%) | 6 (6·0%) | 53 (6·3%) | <6 (4·7%) |
| Different sector | 783 (44·6%) | 54 (6·9%) | 364 (43·5%) | 39 (10·7%) |
| Other  (e.g. parental leave, Sabbatical) | 49 (2·8%) | <6 (1·0%) | 23 (2·8%) | <6 |
| Retired | 615 (35·1%) | 34 (5·5%) | 318 (38·0%) | 48 (15·1%) |
| Missing | 53 (3·0%) | <6 (5·0%) | 17 (2·0%) | <6 (0·9%) |
| **Received flu vaccine 2021/2022** | | | | |
| No | 556 (31·7%) | 35 (6·3%) | 246 (29·4%) | 36 (14·6%) |
| Yes | 1040 (59·3%) | 51 (4·9%) | 500 (63·8%) | 66 (12·4%) |
| Missing | 148 (9·0%) | 14 (8·9%) | 57 (6·8%) | <6 (3·8%) |
| **Received flu vaccine 2022/2023** | | | | |
| No | 461 (26·3%) | 31 (6·7%) | 220 (26·3%) | 30 (13·6%) |
| Yes | 644 (36·7%) | 33 (5·1%) | 391 (46·8%) | 50 (12·8%) |
| Missing | 649 (37·0%) | 36 (5·6%) | 225 (26·9%) | 24(22·6%) |
| **Household with children under 14** | | | | |
| No | 1135 (65·0%) | 54 (4·8%) | 652 (78·0%) | 117 (17·9%) |
| Yes | 247 (14·1%) | 22 (8·9%) | 71 (8·5%) | 14 (19·7%) |
| Missing | 372 (21·1%) | 24 (6·5%) | 113 (13·5%) | 20 (17·7%) |
| **SARS-CoV-2 Nucleocapsid (prior infection)** | | | | |
| negative | 1125 (64·1%) | 60 (5·3%) | 198 (23·7%) | 23 (11·6%) |
| positive | 629 (35·9%) | 40 (6·4%) | 623 (74·5%) | 81 (13·0%) |
| Missing | <6 (0·0%) | <6 (0·0%) | 15 (1·8%) | 2 (1·9%) |
| **Household size** | | | | |
| Median number of household members (IQR) | 2 (2-3) | 2 (2-3) | 2 (2-3) | 2 (2-2) |
| **Household and Non-household contacts** | | | | |
| Median contact number (IQR) | 5 (2-9) | 6 (3-13) | 4 (1-8) | 5 (1-10) |

### 2. Directed Acyclic Graphs

### 2.1. DAG: Association of age and RSV Reinfection risk

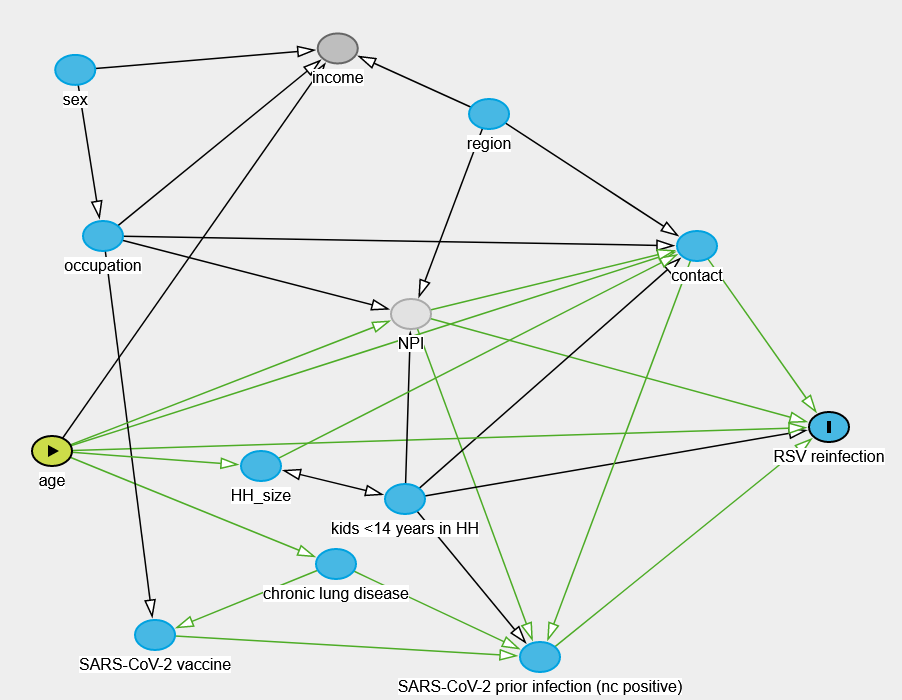

Figure S2: DAG for association of age and RSV Reinfection risk

| Minimal adjustment set | Exposure: age  Outcome: RSV reinfection  No open biasing paths.  No adjustment is necessary to estimate the total effect of age on RSV reinfection. |
| --- | --- |
| Code for DAGitty | dag {  bb="-5.37,-6.752,5.21,6.931"  "RSV reinfection" [outcome,pos="1.799,3.263"]  "SARS-CoV-2 prior infection (nc positive)" [pos="-0.335,6.298"]  "SARS-CoV-2 vaccine" [pos="-3.177,6.007"]  "chronic lung disease" [pos="-1.841,5.071"]  "kids <14 years in HH" [pos="-1.331,4.213"]  HH_size [pos="-2.395,3.777"]  NPI [latent,pos="-1.287,1.772"]  age [exposure,pos="-3.938,3.580"]  contact [pos="0.824,0.875"]  income [pos="-1.828,-1.732"]  occupation [pos="-3.561,0.743"]  region [pos="-0.711,-0.867"]  sex [pos="-3.766,-1.449"]  "SARS-CoV-2 prior infection (nc positive)" -> "RSV reinfection"  "SARS-CoV-2 vaccine" -> "SARS-CoV-2 prior infection (nc positive)"  "chronic lung disease" -> "SARS-CoV-2 prior infection (nc positive)"  "chronic lung disease" -> "SARS-CoV-2 vaccine"  "kids <14 years in HH" -> "RSV reinfection"  "kids <14 years in HH" -> "SARS-CoV-2 prior infection (nc positive)"  "kids <14 years in HH" -> NPI  "kids <14 years in HH" -> contact  "kids <14 years in HH" <-> HH_size  HH_size -> contact  NPI -> "RSV reinfection"  NPI -> "SARS-CoV-2 prior infection (nc positive)"  NPI -> contact  age -> "RSV reinfection"  age -> "chronic lung disease"  age -> HH_size  age -> NPI  age -> contact  age -> income  contact -> "RSV reinfection"  contact -> "SARS-CoV-2 prior infection (nc positive)"  occupation -> "SARS-CoV-2 vaccine"  occupation -> NPI  occupation -> contact  occupation -> income  region -> NPI  region -> contact  region -> income  sex -> income  sex -> occupation  } |

### 2.2. DAG: Association of living with children and RSV Reinfection risk

Figure S3: DAG for kids in HH (<14y) and RSV reinfection risk

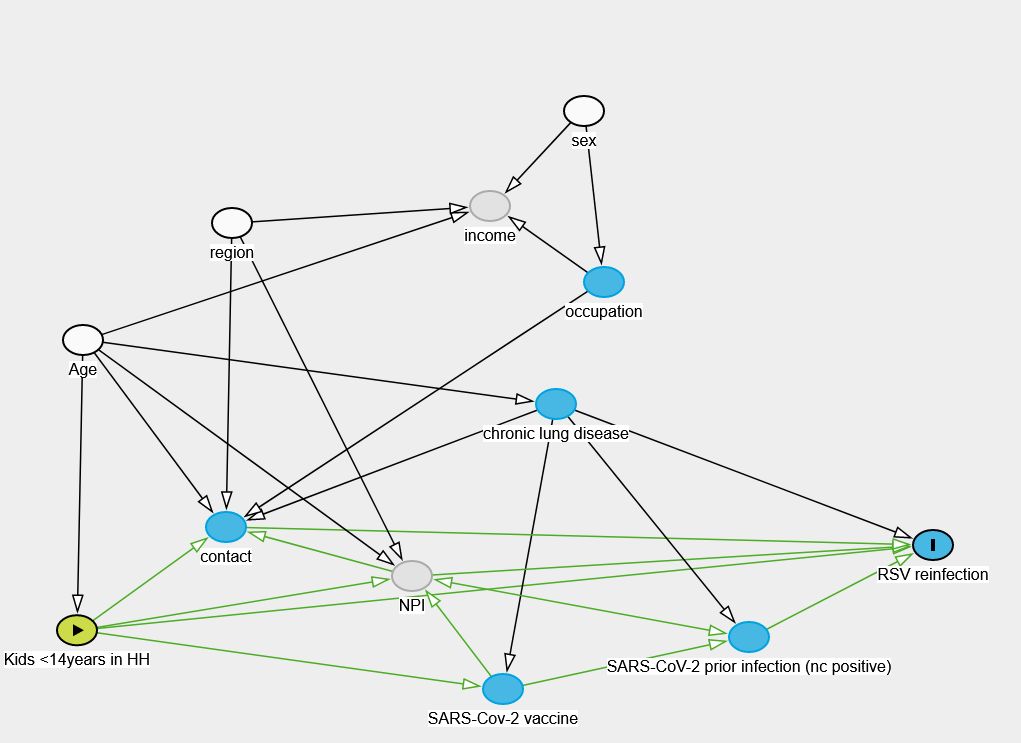

| Minimal adjustment set | Exposure: Kids (<14y) in HH  Exposure: Kids <14years in HH  Outcome: RSV reinfection  Adjusted: Age,region,sex  Correctly adjusted.  Minimal sufficient adjustment sets containing Age, region, sex for estimating the total effect of Kids <14years in HH on RSV reinfection:  Age, region, sex |
| --- | --- |
| Code for DAGitty | dag {  "Kids <14years in HH" [exposure,pos="-1.876,0.983"]  "RSV reinfection" [outcome,pos="-1.240,0.848"]  "SARS-CoV-2 prior infection (nc positive)" [pos="-1.377,0.994"]  "SARS-Cov-2 vaccine" [pos="-1.559,1.076"]  "chronic lung disease" [pos="-1.520,0.625"]  Age [adjusted,pos="-1.872,0.524"]  NPI [latent,pos="-1.627,0.897"]  contact [pos="-1.765,0.820"]  income [latent,pos="-1.569,0.312"]  occupation [pos="-1.484,0.433"]  region [adjusted,pos="-1.761,0.339"]  sex [adjusted,pos="-1.499,0.162"]  "Kids <14years in HH" -> "RSV reinfection"  "Kids <14years in HH" -> "SARS-Cov-2 vaccine"  "Kids <14years in HH" -> NPI  "Kids <14years in HH" -> contact  "SARS-CoV-2 prior infection (nc positive)" -> "RSV reinfection"  "SARS-CoV-2 prior infection (nc positive)" <-> NPI  "SARS-Cov-2 vaccine" -> "SARS-CoV-2 prior infection (nc positive)"  "SARS-Cov-2 vaccine" -> NPI  "chronic lung disease" -> "RSV reinfection"  "chronic lung disease" -> "SARS-CoV-2 prior infection (nc positive)"  "chronic lung disease" -> "SARS-Cov-2 vaccine"  "chronic lung disease" -> contact  Age -> "Kids <14years in HH"  Age -> "chronic lung disease"  Age -> NPI  Age -> contact  Age -> income  NPI -> "RSV reinfection"  NPI -> contact  contact -> "RSV reinfection"  occupation -> contact  occupation -> income  region -> NPI  region -> contact  region -> income  sex -> income  sex -> occupation  } |

### 2.3. DAG: Association of SARS-CoV-2 previous Infection and RSV Reinfection risk

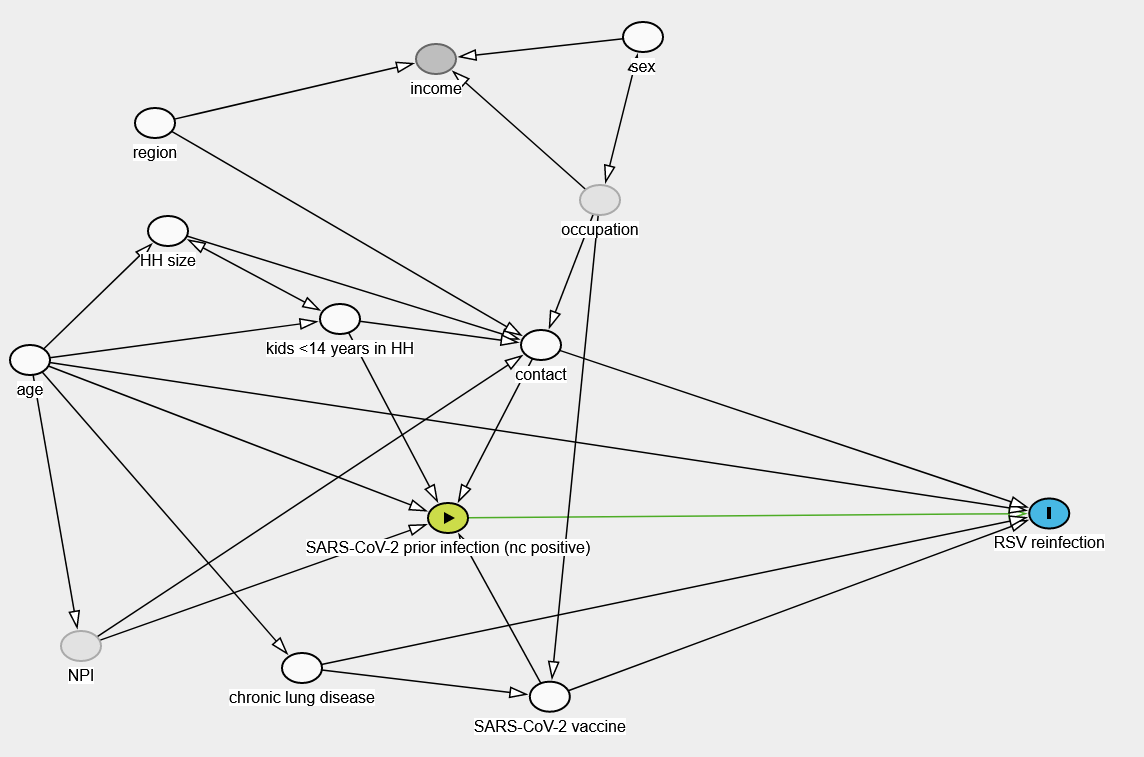

Figure S4: DAG for association of prior SARS-CoV-2 infection and RSV reinfection risk

| Minimal adjustment set | Exposure: SARS-CoV-2 prior infection (nc positive)  Outcome: RSV reinfection  Adjusted: HH size,SARS-CoV-2 vaccine,age,chronic lung disease,contact,kids <14 years in HH,region,sex  Correctly adjusted.  Minimal sufficient adjustment sets containing HH size, SARS-CoV-2 vaccine, age, chronic lung disease, contact, kids <14 years in HH, region, sex for estimating the direct effect of SARS-CoV-2 prior infection (nc positive) on RSV reinfection:  HH size, SARS-CoV-2 vaccine, age, chronic lung disease, contact, kids <14 years in HH, region, sex |
| --- | --- |
| Code for DAGitty | dag {  "HH size" [adjusted,pos="-1.538,0.711"]  "RSV reinfection" [outcome,pos="-0.253,1.224"]  "SARS-CoV-2 prior infection (nc positive)" [exposure,pos="-1.129,1.232"]  "SARS-CoV-2 vaccine" [adjusted,pos="-0.981,1.557"]  "chronic lung disease" [adjusted,pos="-1.342,1.505"]  "kids <14 years in HH" [adjusted,pos="-1.287,0.870"]  NPI [latent,pos="-1.664,1.465"]  age [adjusted,pos="-1.739,0.945"]  contact [adjusted,pos="-0.994,0.918"]  income [pos="-1.147,0.398"]  occupation [latent,pos="-0.908,0.654"]  region [adjusted,pos="-1.557,0.514"]  sex [adjusted,pos="-0.845,0.358"]  "HH size" -> contact  "HH size" <-> "kids <14 years in HH"  "SARS-CoV-2 prior infection (nc positive)" -> "RSV reinfection"  "SARS-CoV-2 vaccine" -> "RSV reinfection"  "SARS-CoV-2 vaccine" -> "SARS-CoV-2 prior infection (nc positive)"  "chronic lung disease" -> "RSV reinfection"  "chronic lung disease" -> "SARS-CoV-2 vaccine"  "kids <14 years in HH" -> "SARS-CoV-2 prior infection (nc positive)"  "kids <14 years in HH" -> contact  NPI -> "SARS-CoV-2 prior infection (nc positive)"  NPI -> contact  age -> "HH size"  age -> "RSV reinfection"  age -> "SARS-CoV-2 prior infection (nc positive)"  age -> "chronic lung disease"  age -> "kids <14 years in HH"  age -> NPI  contact -> "RSV reinfection"  contact -> "SARS-CoV-2 prior infection (nc positive)"  occupation -> "SARS-CoV-2 vaccine"  occupation -> contact  occupation -> income  occupation <-> sex  region -> contact  region -> income  sex -> income  } |

### 3. Logistic regression

The CIs for the RRs were obtained by first deriving the standard errors of the RRs with the Delta-Method from the standard errors of the coefficients in the respective fitted logistic model. In a second step these standard errors were used to construct a CI based on normal approximation. In order to ensure non-negativity of the CI, the standard errors of the logarithm of the RRs (log(RR)) were derived with the Delta-Method and used to construct a CI for log(RR) based on normal approximation. This CI was then in a final step transformed back to the original scale by the exponential function.

Table 2 Multivariable analysis of MuSPAD participant’s characteristics influencing RSV reinfections for season 2020/2021 and 2022/2023

|  | 2020/21-2022 | | 2022-2023 | |
| --- | --- | --- | --- | --- |
| Exposure variables | **RR^1^** | **95% CI** | **RR^1^** | **95% CI** |
| Age group^2^ (reference: 18-29 years ) |  |  |  |  |
| 30-39 | 3.11 | 0.96-10.07 | 0.21 | 0.08- 0.56 |
| 40-49 | 1.88 | 0.56-6.31 | 0.30 | 0.13- 0.68 |
| 50-59 | 0.90 | 0.27-3.01 | 0.24 | 0.12- 0.48 |
| 60-69 | 1.41 | 0.44-4.57 | 0.30 | 0.16- 0.57 |
| 70+ | 1.10 | 0.32-3.69 | 0.39 | 0.20- 0.73 |
| SARS-CoV-2 prior infection (NC)^3^ (reference: NC negative) | 0.65 | 0.33-1.30 | 1.01 | 0.68-1.49 |
| Children in HH<14years^4^ (reference: No) | 1.67 | 0.89-3.13 | 1.32 | 0.88-1.96 |

^1^adjusted relative risk (RR):

^2^Age groups exposition: no adjusted variables for both seasons;

^3^SARS-CoV-2 positive Nucleocapsid exposition adjusted in 2020/21 for HH size, age, chronic lung disease, contacts, kids <14 years in HH, region, sex; in 2022/23 for HH size, age, contact, kids <14 years in HH, region, sex. Adjustment for SARS-CoV-2 vaccine was not possible due to statistical reasons;

^4^Children in HH exposition adjusted for age, region, sex in both seasons;

Further we looked into the number of household and non-household contacts and calculated the Odds ratios (OR with 95% CI) for a 20%- increase of post-F-protein and 45% increase of the N-protein antibodies due to household contacts and non-household contacts.

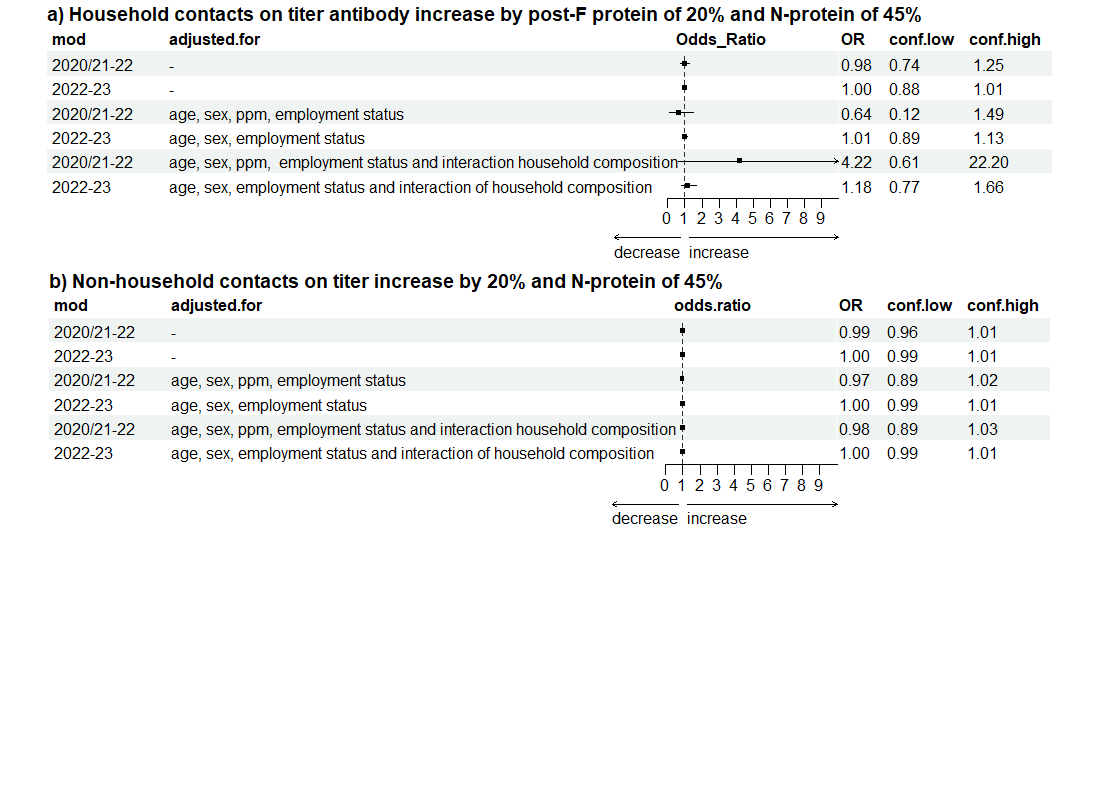

Figure 5: Forestplot of odds ratios (OR with CI-95%) for a 20%- increase of post-F-protein and 40% increase of N-protein attributed to a) household and b) non-household contacts, presented separately for each regression model for each season 2020/21-2022 and 2022/23 and adjustment set (ppm=personal protection measures).

### 4. Data sets for modelling

To estimate the number of RSV cases and RSV hospitalisation rates in the German population we used difference data sources:

*Population data for extrapolation*

We utilized data from Saxony and Germany, obtained from federal state saxony^1^ and the national population statistics^2^ to extrapolate population figures based on age groups and specific years.

*Age-specific RSV notifications*

We use notification data on the number of RSV cases from Saxony (the only German state with RSV reporting until week 40/2023) from 2014 to 2023 from SurvStat@RKI^3^ to estimate extrapolated age-specific RSV case notification data for Germany.

*Acute Respiratory illness (ARI) for age-specific RSV proportions:*

Robert Koch Institute (RKI) collects syndromic data by the sentinel practices of the Arbeitsgemeinschaft Influenza (AGI)^4^ on ARI (acute respiratory illness (cough OR sore throat OR fever) and the subgroup ILI (influenza-like illness, fever AND cough OR sore throat). The RKI reports weekly in form of a report/dashboard.

We integrated RSV diagnoses proportions obtained from weekly acute respiratory infections (ARI) and Influenza-Like Illness (ILI) reports by Arbeitsgemeinschaft (AG) Influenza^5^ (sentinel) and ARI data from GrippeWeb^6^(citizen science surveillance).

Currently, there are approximately 700 medical practices participating, with around 60% of them transmitting electronic data (SEED/ARI). This ARI sentinel network represents roughly 1% of primary care physicians in Germany. Additionally, more than 100 sentinel practices send patient samples to the RKI. The national reference center (NRC) examines these to determine currently circulating respiratory viruses.

RKI reports the number of patients with ARI who visit physician's offices per 100,000 inhabitants within specific age groups (divided into five age groups 0-4, 5-14, 15-34, 35-59, 60+). Nationwide data has been available since the 2012/13 season. It is important to note that information on the proportion of RSV infections is limited, with data accessible for the 2016/2017 season, weeks 1-14 in 2018, weeks 4-14 and 49 in 2019, no RSV data for 2020, weeks 10-20 in 2021, and the 2021/2022 and 2022/2023 seasons.

The ARI data are available under:

- <https://github.com/robert-koch-institut/ARE-Konsultationsinzidenz>
- and the reports with RSV proportion https://influenza.rki.de/Wochenberichte.aspx.

We calculated **age-specific RSV proportions from sentinel data** for application to Grippe-Web.

*GrippeWeb (citizen science) for RSV infections in GrippeWeb*

GrippeWeb is an online syndromic monitoring system for tracking acute respiratory illness in the general population of Germany. This system was initiated in 2011. Registered participants who are 14 years or older have the option to complete a baseline survey that includes demographic information such as age, place of residence, and past illnesses. Additionally, weekly questionnaires are distributed, inquiring about new respiratory illnesses, symptoms experienced, and whether a medical professional was consulted. Furthermore, participants have the option to include their children in the system and respond to questions regarding respiratory illnesses on their behalf, including Acute Respiratory Illness (ARI) and Influenza-Like Illness (ILI) as defined. Grippe-Web data (5,878 GrippeWeb-participants as of 41/2023) are available in the aggregated age groups (0-5, 6-14, 15-24, 25-39, 40-59, 60+) and can be downloaded: <https://github.com/robert-koch-institut/GrippeWeb_Daten_des_Wochenberichts/blob/main/GrippeWeb_Daten_des_Wochenberichts.tsv>. We estimated **RSV infections in GrippeWeb**.

*Severe Acute Respiratory Infections (SARI) for Proportion of RSV-related hospitalizations*

Given that RSV represents a substantial disease burden and is a primary contributor to hospital admissions due to SARI among children aged ≤2 years and >65 years in Germany, we utilized RKI data on the incidence of cases admitted to hospitals for SARI to estimate the RSV **proportion of hospitalized cases** associated with severe symptomatic respiratory infections. This calculation relies on data obtained from the ICOSARI syndromic hospital surveillance system (representing 6-7% of the German hospitals).^7^ This sentinel surveillance started collecting data with season 2015/16. Only for season 2022/23 the proportion of RSV hospitalization is accessible.

Data available under: <https://github.com/robert-koch-institut/SARI-Hospitalisierungsinzidenz/blob/main/SARI-Hospitalisierungsinzidenz.tsv>

More information on SARI Surveillance: <https://www.rki.de/DE/Content/Infekt/Sentinel/SARI-KH-Sentinel/node.html>

This pyramid figure 1 depicts the acute respiratory illness of the Sentinel-Surveillance system by the RKI.

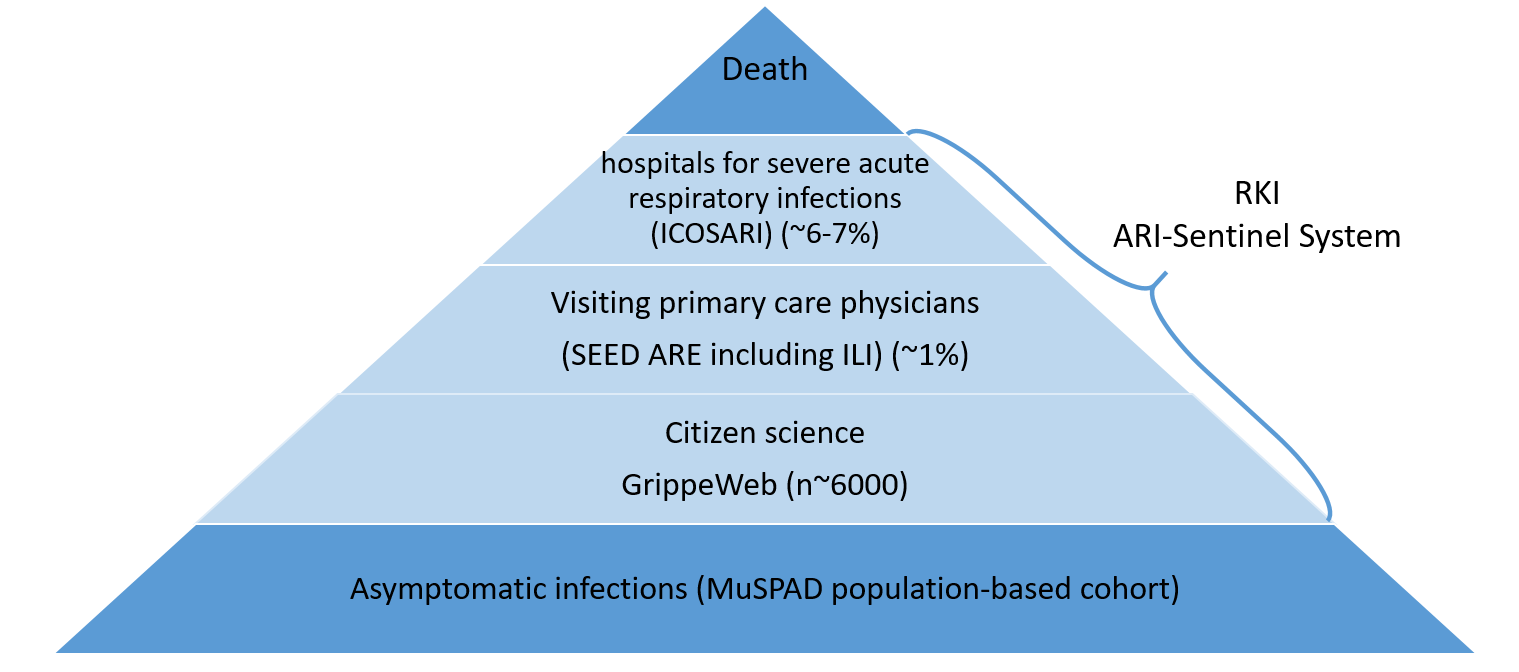

Figure S6: Sentinel and surveillance data sources for RSV in Germany^8^; modified by us

### 5. Calculations

*RSV related infections in Germany (Notification and sentinel data ARI)*

To calculate the number of RSV infections, we used two distinct methods, one with notification data (1) and one with sentinel data (2).

In the method (1), we downloaded the RSV infection numbers in Saxony from SurvStat@RKI and extrapolated the data on the population of Germany based on the number of population.

In the method (2), we took the ARI numbers reported in the GrippeWeb as basis for the number of infections. Since the true proportion of RSV in the ARI rates is unknown, we used the proportion of cases tested positive on RSV (RSVprop.Sent.surv.) from the sentinel surveillance reported by Influenza AG (NRC) in the weekly reports to subset the ARI rates (equation eq. 1.1).

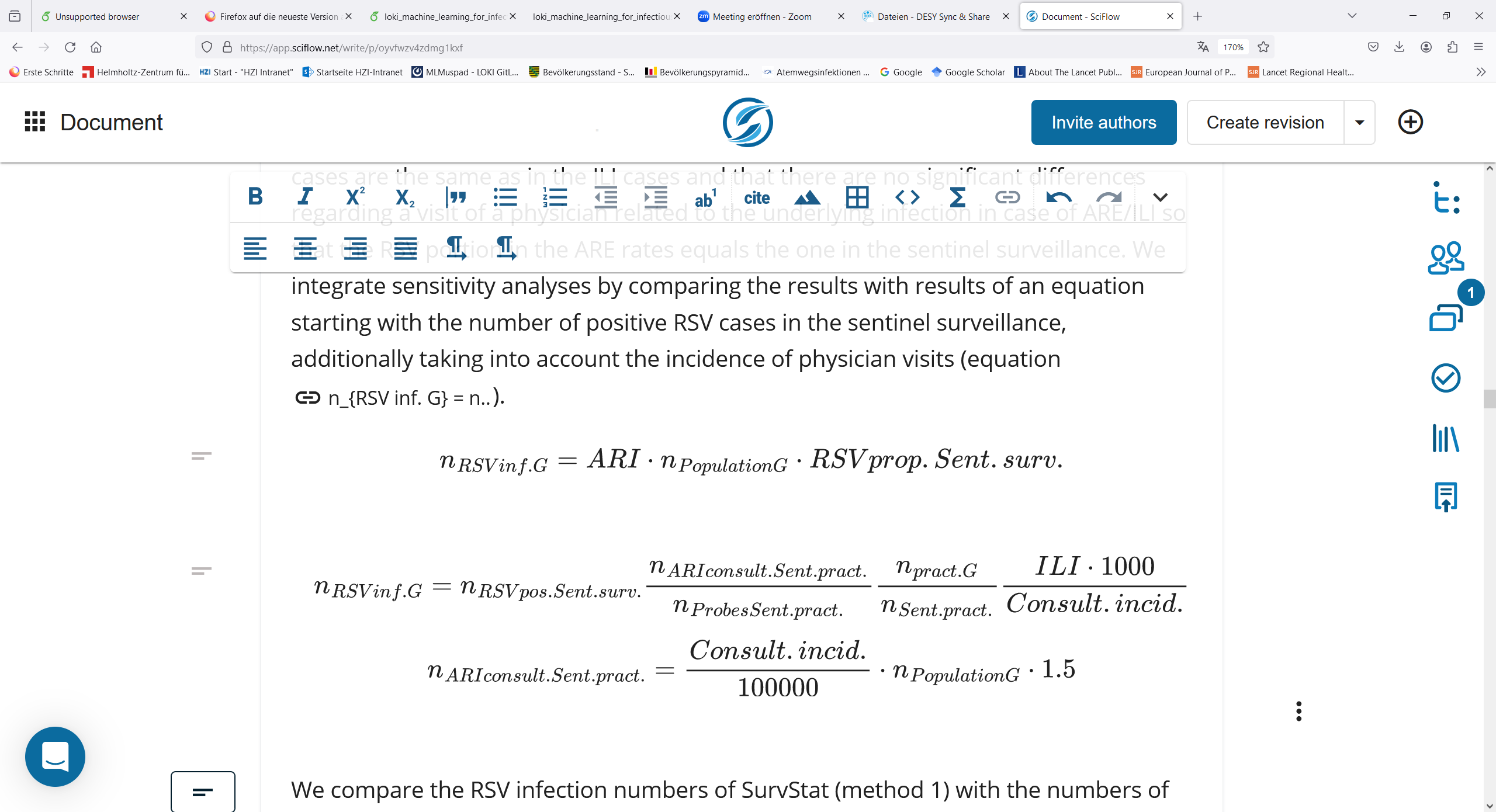

The outcome is an estimation of the number of RSV infections in Germany (n_RSVinf.G_) stratified for age groups over the calendar weeks. We assume that the proportion of persons being tested with RSV in the ARI cases are the same as in the ILI cases. We also assume that there are no notable variations in the likelihood of seeking medical care related to the underlying infection in cases of ARI or ILI. Consequently, the proportion of RSV in the ARI rates matches the one observed in the sentinel surveillance data. (SEED ARI). We integrated sensitivity analyses by comparing the results with results of an equation starting with the number of positive RSV cases in the sentinel surveillance ($n_{RSVpos.Sent.surv.}$), additionally taking into account the incidence of physician visits (equation eq. 1.2). Hereby, the number of positive RSV cases in the sentinel surveillance ($n_{RSVpos.Sent.surv.}$) is extrapolated from the people being swabbed ($n_{ProbesSent.prect.}$) to all people visiting the Sentinel clinics with ARI symptoms ($n_{ARIconsult.Sent.pract.}$). Then it is extrapolated to the number of people visiting all german clinics with ARI symptoms (dividing by the number of sentinel clinics, $n_{Sent.pract.} ,$and multiplying with the total number of clinics in Germany, $n_{pract.G}$).  At last it is extrapolated from the people consulting a clinic with ARI or ILI to all people with ARI/ILI symptoms in Germany (since the consultation incidence is a number per 100000 and ILI/ARI are given per 100, latter proportion is multiplied by 1000).

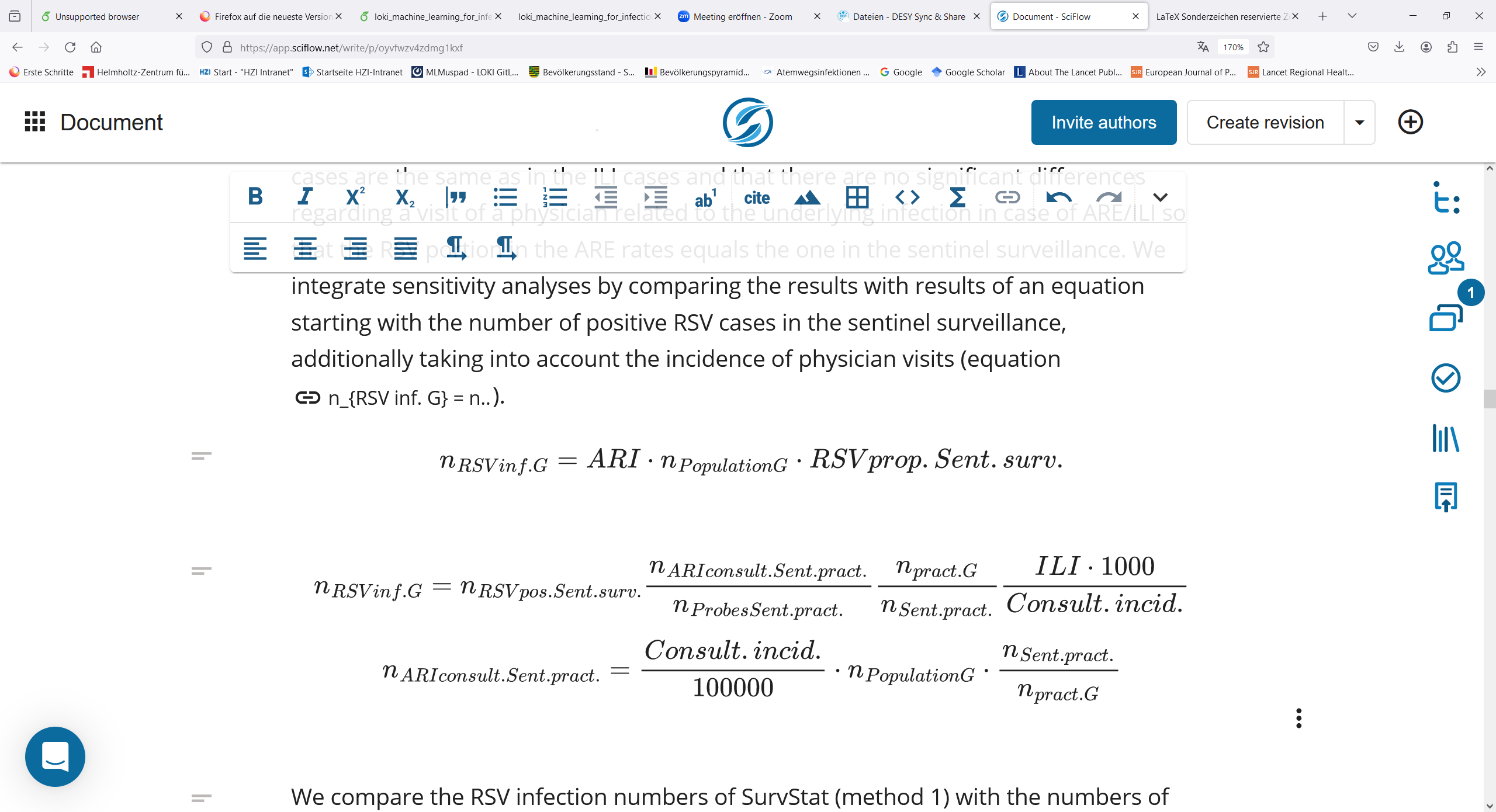

We compared the RSV infection numbers of SurvStat@RKI (method 1) with the numbers of the ARI sentinel systems with RSV proportion (method 2) to estimate underdetection factors for each age groups of the mandatory reporting system over time.

*Calculation of underdetection factor:*

We calculated an underdetection ratio by dividing the RSV number ARE sentinel system by SurvStat@RKi RSV cases extrapolated to Germany (and we separately include the hospitalization statistics) for each age group across calendar weeks. This allows us to compare RSV hospitalizations and infections.

*RSV related hospitalisation in Germany (SARI)*

We generate an indicator of the true number of infections and hospitalizations in each age groups over time, including an indicator for susceptible children. To calculate the hospitalization rates, we use the RSV proportion of the Hospital sentinel ICOSARI (ICD-10 code-based hospital surveillance of severe acute respiratory infections) surveillance from the Influenza AG. Since it reports the number of positive tested RSV cases in the sentinel hospitals, we expand it to all German (G) hospitals by the following equation eq. 1.3

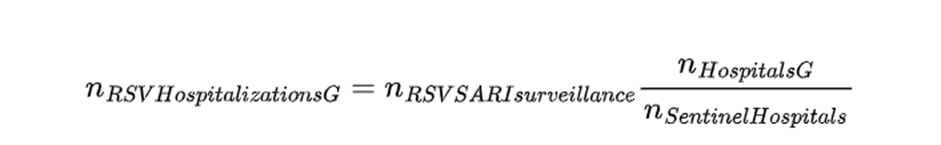
 We assume that the proportion of hospitalized patients is consistent for Influenza-Like Illness (ILI) across Germany. Therefore, we are able to extrapolate the data from the sentinel hospitals to encompass all hospitals in Germany.

### 6. Age-specific RSV infections

The detailed equations of model are following.

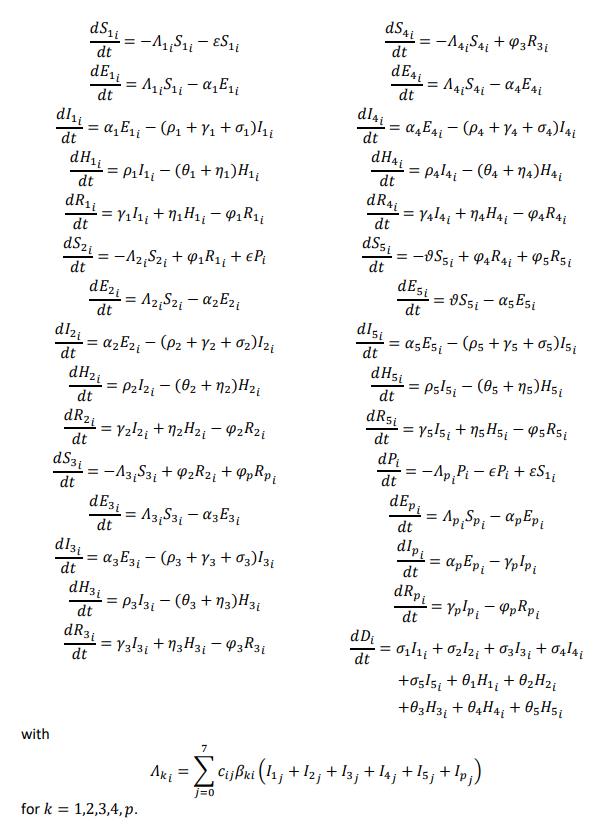

Table 2: Parameterization of ODE model for RSV infection

| Parameter | Description | Reference |
| --- | --- | --- |
| Infection risk | Risk of infection per contact | Model fitting |
| Contact rate | Average number of contact per person per day | COVIMOD^9^ |
| Incubation period | Time from infection to onset of symptoms | Model assumption (RKI)^10^ |
| Infectious period | Duration a person is infectious | Model assumption (ECDC)^11^ |
| Hospitalization rate | Proportion of infected individuals requiring hospitalization | Model fitting |
| Recovery rate | Rate at which infected individuals recover | Model assumption (ECDC^11^) |
| Mortality rate | Rate at which infected individuals die | Model assumption (CDC^12^) |
| Reinfection rate | Proportion of recovered individuals getting reinfection | Model fitting & MuSPAD |

Figure 6 displays extrapolated **RSV notifications** (extrapolated SurvStat@RKI data) and **SARI RSV hospitalisations** for Germany by age group from 2016-2023. Figure 7 provides the distribution of **SARI RSV hospitalisations** with data from **the citizen science (Grippe Web) and sentinel surveillance (ARI)** over the same period. In both approaches, the highest case numbers are estimated in children under 4 years in 2021 and 2022, followed by 60+ and 35-59 years. The mean hospitalisation rate was about 335 % based on official passive surveillance system and 24 % in our calculated assumed cases. Figure 5 shows the extrapolated SurvStat@RKI data in comparison to the estimated RSV infections based on the other sentinel systems. From 2016-2023 all three different data sources mapped well the seasonal RSV dynamic, for the 2020/2021 season, none of the three figures displayed any RSV wave. When comparing the seasons, we observe a shift in 2021 towards an earlier onset of RSV infections in the fall, along with a higher peak and a more rapid decline in cases according to the SurvStat@RKI data and a delay within the RSV sentinel surveillance (Figure 6). During the 2022/23 season, there is an improved alignment between the two data sources for the age group 0-4 years.

The average estimated underreporting ratio is 86.2 when considering all age groups and weeks. The mean underreporting ratio was highest in 2016 followed by 2017 and 2022. Within the age group 35-59 years, a substantial proportion of infections went unreported in the passive surveillance system, especially in 2027 and 2022. In comparison to the other years, the underreporting rate was at its lowest in 2023. We found two extremely high peaks of underreporting followed by very low factors that were removed as we assumed a delay in reporting in these cases.

According to the official passive surveillance report of Saxony, 2627 children that are now 0 to 2 year old had an RSV-infection within the last 1.5 years. Assuming an age-specific average underdetection of about 95 % for 2022, the estimated number is 52540 0-2-year-old children. The population number of 0-2-year-old children in Sachsen is 62903, deriving to 84 % of 0-2-year-old children not susceptible to first infection and 16 % (n=10064) still susceptible to first infection.

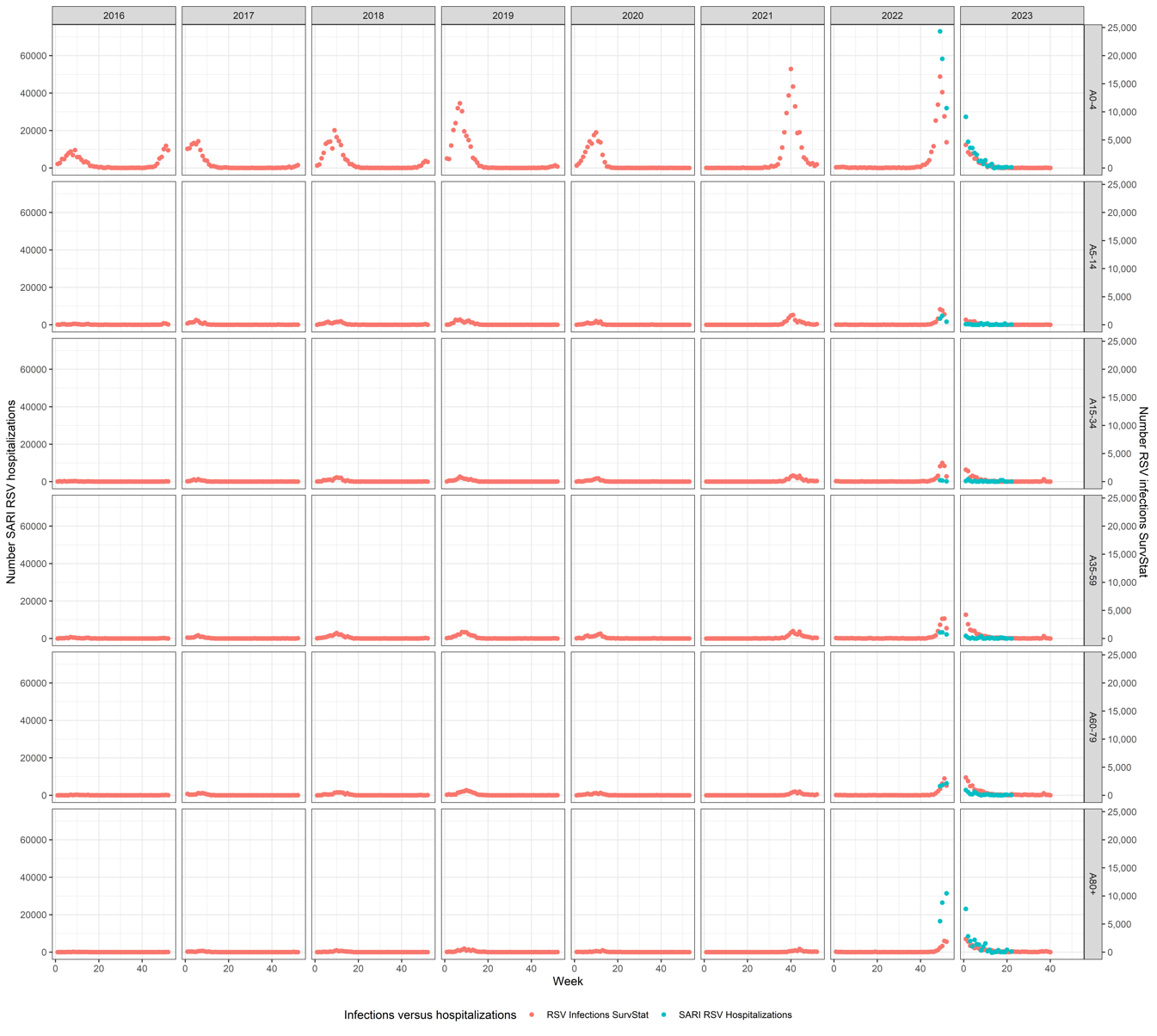

Figure S7: Numbers of RSV-infections from official notification reporting system SurvStat@RKI of Saxony expanded to Germany (red points) and RSV SARI hospitalizations (blue points) in the population from 2016 to 2023 (note: different y axes scaling)

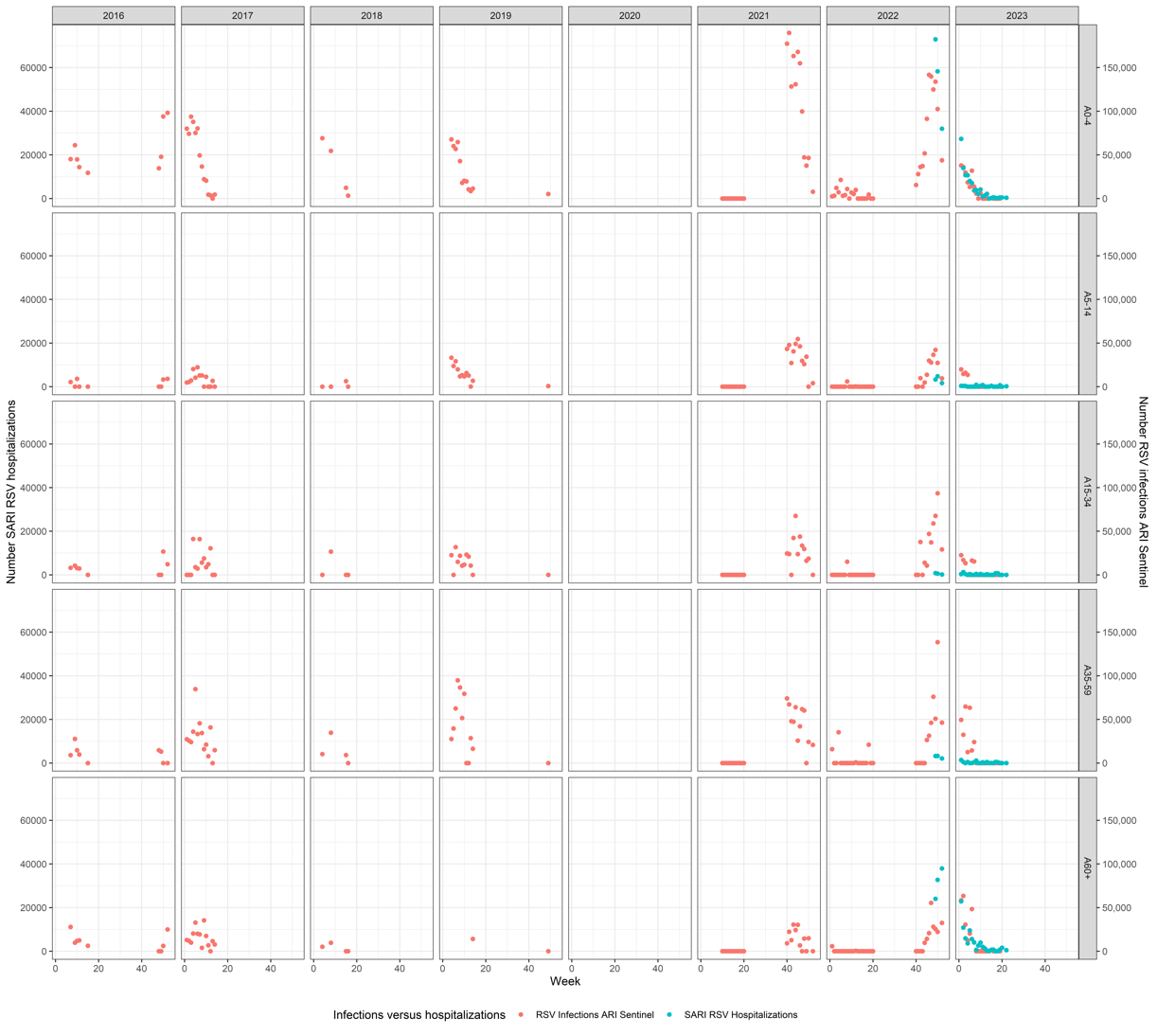

Figure S8: Numbers of RSV-infections based on GrippeWeb and RSV proportions of the ARI sentinel surveillance (red points) in comparison to SARI RSV hospitalizations (blue points) in the German population from 2016 to 2023 (note: different x axes scaling)

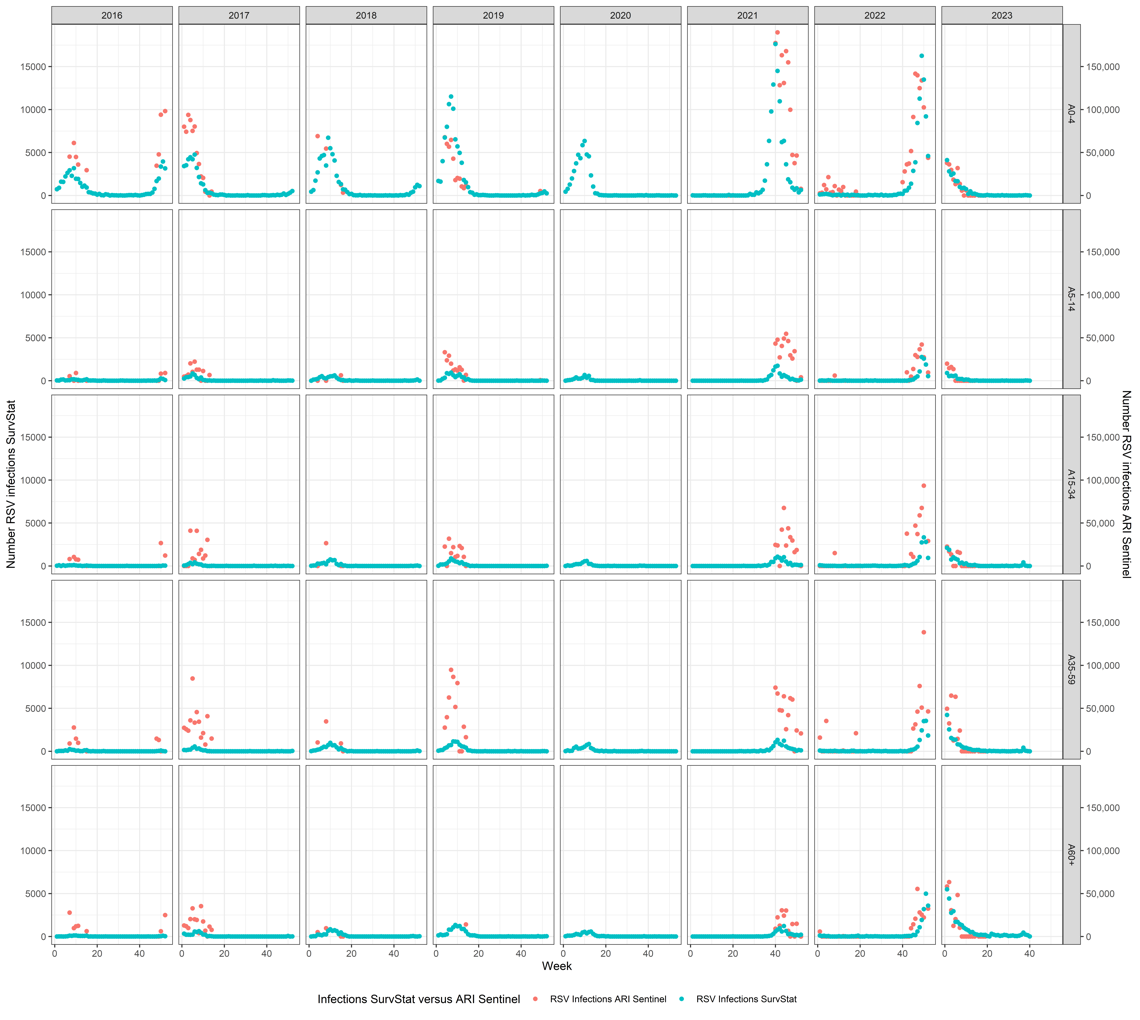

Figure 9: RSV notifications from official notification reporting system SurvStat@RKI of Saxony expanded to Germany (blue points) and estimated cases based on GrippeWeb and the RSV proportions of the ARI sentinel surveillance (red points) from 2016 to 2 to 2023 (note: different x axes scaling and no sentinel data are available for 2020)

### 7. Titre increases indicative of RSV reinfection in the last year

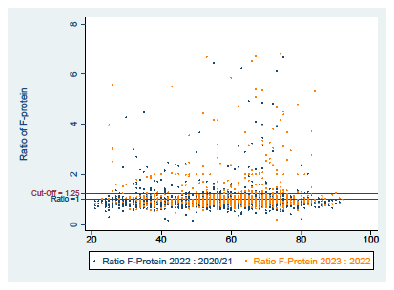

Figure S10: Figure S9: Scatter plot illustrating the ratio of F-Protein for the seasons 2020/21-2022 and 2022-2023 across different age groups. The maroon line on the plot represents cut-off value (25% titre increase) for RSV reinfection. The blue dots on the graph correspond to the season 2020/21-2022, while the orange dots represent the season 2022-2023

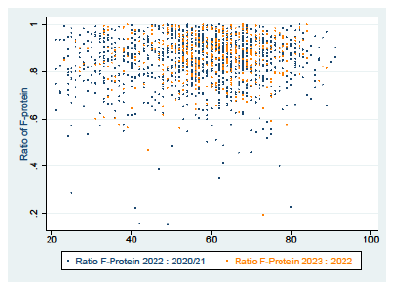

Figure S11: Scatter plot illustrating the ratio of F-Protein for the seasons 2020/21-2022 and 2022-2023 across different age groups if the ratio is below one. The blue dots on the graph correspond to the season 2020/21-2022, while the orange dots represent the season 2022-2023.

### 8. Sensitivity analyses for different cut-offs of the multiplex serology

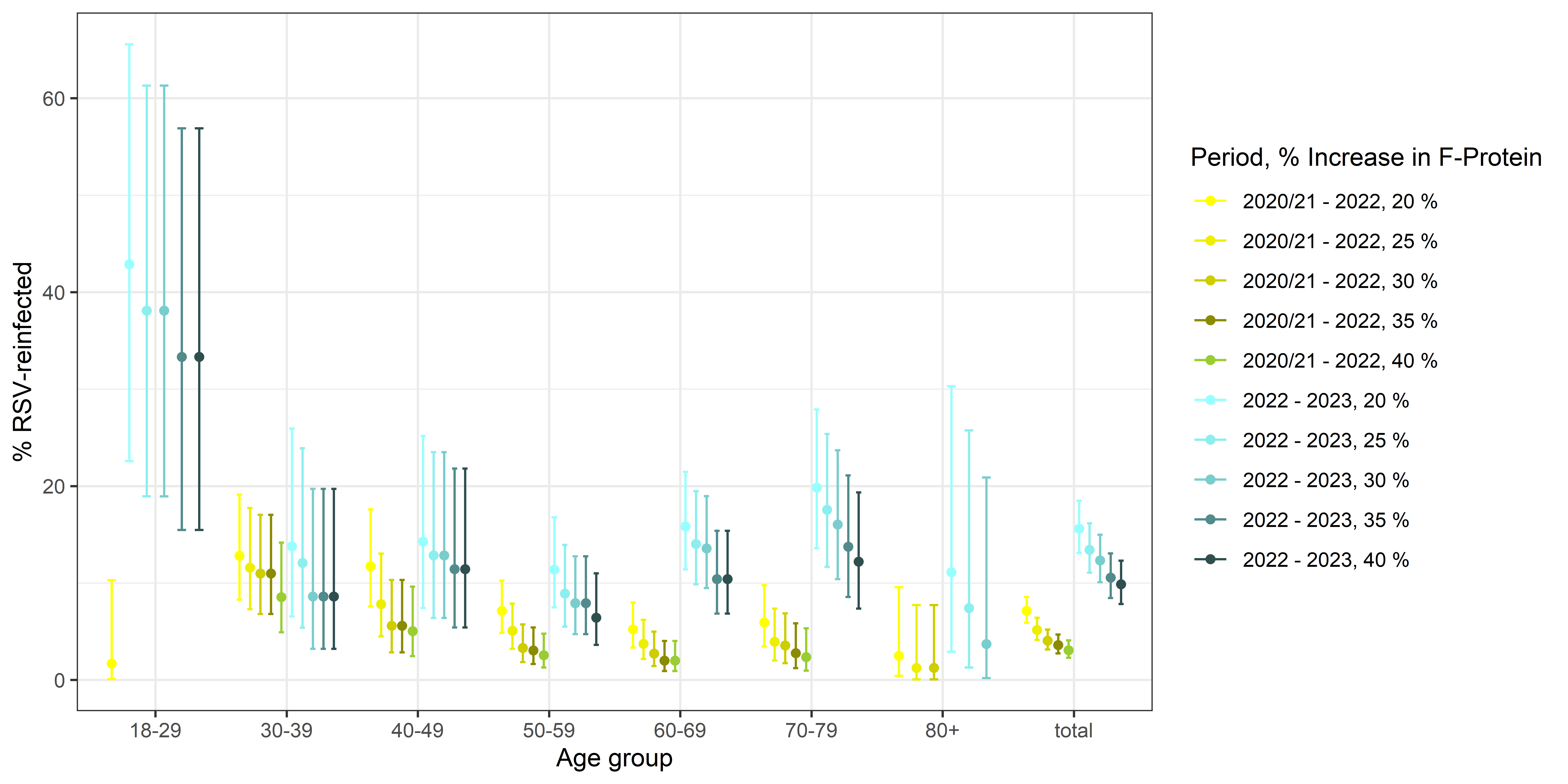

Figure 12: RSV reinfection applying varying RSV titre increases (20%-40%) by age groups in 2020/2021 to 2022 (n=110 out of 1754) and 2023 (n=151 out of 836) in RESPINOW

### 9. References

1.	Sachsen SLdF. Die Datenbank des Statistischen Landesamtes des Freistaates Sachsen. 2023. <https://www.statistik.sachsen.de/genonline/online#astructure> (accessed 01.10. 2023).

2.	Statistisches Bundesamt (Destatis). 12111-0002: Bevölkerung (Zensus): Deutschland, Stichtag, Nationalität, Altersgruppen. 2023. <https://www-genesis.destatis.de/genesis//online?operation=table&code=12111-0002&bypass=true&levelindex=0&levelid=1701766835493#abreadcrumb> (accessed 01.10.2023 2023).

3.	Robert-Koch-Institut. RSV notifications for Saxony. 2021 (accessed accessed 09/2021.

4.	Goerlitz L, Tolksdorf K, Buchholz U, et al. Monitoring of COVID-19 by extending existing surveillance for acute respiratory infections. *Bundesgesundheitsblatt-Gesundheitsforschung-Gesundheitsschutz* 2021; **64**: 395-402.

5.	Dürrwald R, Wedde M, Duwe S, et al. Synopse virologischer Analysen im Nationalen Referenzzentrum für Influenzaviren während der COVID-19-Pandemie. 2022.

6.	Robert-Koch-Institut. GrippeWeb. 2024. <https://www.rki.de/DE/Content/Infekt/Sentinel/Grippeweb/grippeweb_node.html2024>).

7.	Buda S, Tolksdorf K, Schuler E, Kuhlen R, Haas W. Establishing an ICD-10 code based SARI-surveillance in Germany–description of the system and first results from five recent influenza seasons. *BMC public health* 2017; **17**(1): 1-13.

8.	Robert-Koch-Institut. Surveillance-Systeme des RKI zu ARI in Deutschland. 2024. <https://public.data.rki.de/t/public/views/ARE-Dashboard/Ueberblick?%3Aembed=y&%3AisGuestRedirectFromVizportal=y>

9.	Tomori DV, Rübsamen N, Berger T, et al. Individual social contact data and population mobility data as early markers of SARS-CoV-2 transmission dynamics during the first wave in Germany—an analysis based on the COVIMOD study. *BMC medicine* 2021; **19**: 1-13.

10.	Robert-Koch-Institut. RSV-Infektionen RKI-Ratgeber. 2024. <https://www.rki.de/DE/Content/Infekt/EpidBull/Merkblaetter/Ratgeber_RSV.html#doc2394298bodyText62024>).

11.	ECDC. Respiratory syncytial virus (RSV). 2024. <https://www.ecdc.europa.eu/en/respiratory-syncytial-virus-rsv>.

12.	Havers FP. Characteristics and outcomes among adults aged≥ 60 years hospitalized with laboratory-confirmed respiratory syncytial virus—RSV-NET, 12 states, July 2022–June 2023. *MMWR Morbidity and Mortality Weekly Report* 2023; **72**.
